# 2010 and 2013 incidence peaks in narcolepsy and idiopathic hypersomnia linked to type A H1N1 and type B Victoria influenza strains

**DOI:** 10.1101/2024.04.19.24304365

**Authors:** Zhongxing Zhang, Jari K. Gool, Pavel Sirotkin, Yves Dauvilliers, Lucie Barateau, Giuseppe Plazzi, Fabio Pizza, Francesco Biscarini, Karel Sonka, Karolina Galuskova, Aleksandra Wierzbicka, Birgit Högl, Eva Feketeova, Rafael Del Río Villegas, Rolf Fronczek, Gert Jan Lammers, Ramin Khatami

## Abstract

**Introduction:** Increased narcolepsy type 1 (NT1) incidence rates have been reported globally in 2010, and were linked to the type A H1N1 2009-2010 influenza pandemic and Pandemrix vaccination. A European child-specific NT1 incidence peak was additionally observed in 2013 post the H1N1 pandemic. Thus, the relationship between NT1 and influenza infection remains unclear. Whether other influenza viruses may also trigger NT1 or other central disorders of hypersomnolence (CDH), is unknown. This study investigated annual European incidence patterns of all CDH in complete samples from multiple European centers, in relation to the severity of individual flu strains in preceding influenza seasons.

**Methods:** Incidence rates of NT1 (N=981) and the combined group of narcolepsy type 2 (NT2) and idiopathic hypersomnia (IH) (N=545) from eight European countries were temporally analysed to identify possible incidence peaks from 1995 to 2019. Linear mixed models and spearman correlations were conducted between hypersomnolence disorder incidence rates and the number of influenza infections of preceding influenza season, split for types A H1N1 and H3N2, and in the Netherlands also types B Victoria and Yamagata influenza.

**Results:** 2010 and 2013 incidence peaks were present in NT1, and a 2010 children peak was unexpectedly found in the combined group of NT2 and IH. Both hypersomnolence groups exhibited a significantly positive relationship with preceding H1N1 influenza season severity and a negative relationship with H3N2 influenza. NT1 was additionally significantly positively correlated with influenza type B Victoria in the Netherlands and showed highest correlation in children.

**Conclusions:** Besides H1N1 influenza, the temporal association and severity correlation suggest that influenza type B Victoria may be a novel potential trigger for NT1 that requires further investigation. We additionally provide insights into possible immune-related pathophysiologies of NT2 and IH associated with the 2009-2010 H1N1 influenza pandemic. Further immunological investigations are warranted to unravel the complexities of these relationships and their implications for CDH.

## Introduction

Narcolepsy type 1 (NT1) is a neurological disorder characterized by excessive daytime sleepiness (EDS) and cataplexy, resulting from the loss or dysfunctioning of hypocretin (or orexin) neurons^1^. A connection with environmental events triggering NT1 gained interest after increased incidence rates were reported in children in North European countries with widespread use of Pandemrix (GlaxoSmithKline Biologicals, Wavre, Belgium), a type A H1N1 influenza vaccine, during the 2009-2010 type A H1N1 influenza pandemic (pH1N1)^2,3^. Subsequent data from France suggested that the use of Pandemirix was associated with increased NT1 incidence rates in both children and adults^4^. Also in East Asia and the United States where Pandemrix was not used, incidence increases were noted, leading to the hypothesis that the type A H1N1 influenza virus itself may also trigger NT1^5–9^. Later genetic evidence further supported this hypothesis^10^. It seems plausible that various triggers must already have been present before the recirculation of type A H1N1 in 2009. Streptococcus pyogenes infections have been associated with onset of NT1 but supporting evidence remains limited^11–15^. The concept of the multiple-hit hypothesis emerged, suggesting that immune-mediated narcolepsy might not be specific to type A H1N1 influenza infection or Pandemrix vaccination but involves various triggers, potentially combined, to induce NT1 in genetically susceptible individuals^16,17^.

Within multiple European countries, a second NT1 incidence peak was discovered for 2013 using the European Narcolepsy Network (EU-NN) database^18^. Most interestingly, this peak was child-specific. These fluctuating narcolepsy incidence rates suggest a temporal relationship between narcolepsy development and environmental fluctuations, such as influenza type and season severity. Influenza seasons differ heavily geographically and on an annual basis in regards to duration, severity and dominance of particular influenza strains. Since the recirculation of type A H1N1 after the pH1N1, four regularly circulating influenza strains are generally identified, type A H3N2, type A H1N1, type B Victoria and type B Yamagata. Where reassortment in type A influenza is much faster, type B influenza generally affects younger individuals and less frequently requires medical consultation^19,20^.

In the United States and China, NT1 incidence rates were only weakly correlated to type A H1N1 influenza season severity^9,21^. This could be driven by geographic differences within these large countries in influenza season severity and strain dominance, or the influence of other, still unknown triggers. Whether the temporal relationship between type A H1N1 influenza season severity and NT1 incidence holds true for Europe and whether other influenza strains could also be linked to NT1 onset, remains to be investigated. Most interestingly, among the dominant influenza strains in the 2012-2013 influenza season (preceding the 2013 NT1 peak in children) was type B influenza in many European countries^22^. This suggests that type B influenza could also be a potential trigger for narcolepsy^18^. Previous analyses only focused on onset of NT1 and lacked inclusion of other central disorders of hypersomnolence (CDH). Narcolepsy type 2 (NT2) and idiopathic hypersomnia (IH) share the complaint of EDS but in the absence of cataplexy and without hypocretin deficit in the cerebrospinal fluid (CSF). The pathophysiologies of these disorders remain elusive but recent findings have suggested frequent reports of respiratory infections close to onset of NT2 and IH^23^. A possible relationship with influenza infections has not yet been carefully studied.

The main aim of the current study was to first investigate NT1, NT2 and IH incidence rates between 2010-2019 (following the pH1N1) using complete samples from multiple European sleep-wake clinics. We additionally correlated incidence rates to detailed yearly influenza season severity data within each country, while taking into account distributions of different influenza strains. Type A influenza was included for all countries, and type B influenza data was only available for the Netherlands. We hypothesized a significant correlation between type A H1N1 and possibly type B influenza and NT1 incidence rates in children.

## Methods

All people with CDH who attended one of our specialized sleep-wake clinics between January 2010 and December 2019 were included in this observational study. Clinics included Sleep-Wake Center SEIN in Heemstede (the Netherlands, total sample is N=295), Montpellier (France, N=619), Bologna (Italy, N=246), Prague (Czech Republic, N=156), Warsaw (Poland, N=94), Kosice (Slovak Republic, N=37), Innsbruck (Austria, N=45) and Madrid (Spain, N=34). Medical records were retrospectively reviewed and detailed data were extracted on the symptoms (EDS and cataplexy presence, and EDS date of onset and Epworth Sleepiness Scale score, ESS), sleep test results (multiple sleep latency test [MSLT] mean sleep latency and sleep-onset rapid eye movement period [SOREMP] count and polysomnography SOREMP presence) and objective biomarkers (CSF hypocretin-1 levels, and HLA-DQB1*06:02 positivity). Hypersomnolence diagnoses were compliant with ICSD-3 criteria and individuals were classified as having NT1 or NT2, or IH^24^. Centers generally performed a diagnostic protocol in which nocturnal polysomnography was directly followed by a MSLT. In Montpellier, the 32-hour bed rest protocol was used for diagnosing IH in case of long sleep time^25^. In our incidence rate and influenza season correlational analyses we merged the individuals with NT2/IH into one group to achieve sufficient power. Those with a clear complaint of EDS almost fulfilling ICSD-3 diagnostic criteria were included as people with a “clinical diagnosis” (for instance a person with typical cataplexy, MSLT sleep latency of four minutes and one SOREMP). Supplementary material 2 shows an overview of the clinical diagnosis groups’ definitions, number of subjects per center, and repeated sensitivity analyses without the people with a clinical diagnosis to investigate whether this group had substantially influenced our results.

Demographics and diagnostic criteria were statistically compared between NT1, NT2 and IH. Shapiro–Wilk normality tests were first used to test the distributions of the numerical variables in each subgroup, and one-way ANOVA or Kruskal-Wallis tests were used to compare the variables among the three subgroups depending on whether the data were normally distributed.

All sites are members of EUNN and they provided pseudonymised data with ethical approval from their local ethics committees and institutional review boards (i.e., the Netherlands: the Medical Ethical Committee of the VU Medical Center scrutinized the study as it consisted of an analysis of previously acquired clinical data posing no risk to included individuals [reference number: 2020.109]; Montpellier, France: Comité de Protection des Personnes France [reference number: 018-A00703-52]; Bologna, Italy: Comitato Etico di Area Vasta Emilia Centro [reference number: EM539-2022-17009-EM1-OSS-AUSLBO]; Prague, Czech Republic: Etická komise Všeobecné fakultní nemocnice v Praze [reference number: 115/21 S]; Warsaw, Poland: Instytut Psychiatrii I Neurologii Komisja Bioetyczna [reference number 21/2010]; Košice, Slovak Republic: Etická komisia Univerzitnej nemocnice L. Pasteura Košice [reference number 22.05.2014]; Innsbruck, Austria: Eithikkommission der Medizinischen Universität Innsbruck [reference number: AH3368 269/4.7 389/5.13(4311a)]; Madrid, Spain: Comité Ético de Investigación Clínica del Grupo Hospital de Madrid [reference number: 15.02.748-GHM]). The clinical experiments conformed to the principles outlined by the Declaration of Helsinki.

### Disease onset

We chose the year of EDS onset as disease onset, as it in general is the first symptom of narcolepsy or IH to develop. In case of unclear onset, for instance when an individual was unsure about onset between two years, this individual was partitioned in the incidence rates for both years. Subjects with unclear disease onset were excluded from our analyses.

### Hypersomnolence incidence rates

We first repeated the locally estimated scatterplot smoothing (LOESS) models as described in Zhang et al.^18^. We analysed NT1 (N = 981) separately from the combined group of NT2 (N =189) and IH (N = 356). In summary, the incidence rate of each year was predicted by the LOESS model with 95% confidence intervals (CIs) and then the ratio R was calculated between the predicted value and the observed value in each year. LOESS models were first performed combining data from all countries and hereafter repeated separately in the countries with at least 150 individuals (the Netherlands, France, Italy and Czech Republic). The incidence rates were considered as R-fold significantly increased if the bottom of the 95% predictive CIs of R was larger than 1.3 and the P-value < 0.0001. Sensitivity analyses were performed for the combined group of NT2/IH to test the robustness of possible incidence peaks. To test whether HLA-DQB1*06:02 positivity may have played a role, we first excluded the individuals that were HLA-DQB1*06:02 positive. We additionally implemented permutation testing in which we repeated the LOESS model 10,000 times, each with a 90% randomly sampled proportion of the children with NT2/IH. As an exploratory post-hoc analysis we also repeated the incidence rate analyses split for NT2 and IH in Supplementary material 1.

In addition, we divided the database into two subgroups: children cases (age of EDS onset ≤ 18 years old) and adult cases (age of EDS onset > 18 years old), and repeated the LOESS modelling in the two subgroups to further investigate whether possible increased incidence rates of the hypersomnolence disorders were age-specific. Age of symptom onset was missing in for some individuals in the databases from Innsbruck (29 NT1 and 16 IH with missing onset), Czech Republic (1 NT1) and the Slovak Republic (2 NT1) and these people were thus excluded from the analyses separating child- and adult-onset.

### Influenza season severity correlations

For the countries with largest hypersomnolence incidence datasets (the Netherlands, France, Italy and Czech Republic, which also included data of both NT1 and NT2/IH patients) we additionally performed correlational analyses between hypersomnolence disorder incidence rates and influenza season severity defined by the number of infections in the preceding season (e.g., between the influenza season severity in 2009-2010 and narcolepsy incidence in 2010) with additional analyses on the different influenza strains. We again separately analysed NT1 from the combined group of NT2/IH. Detailed yearly influenza data was extracted for each included country from the *Global Influenza Programme* of the *World Health Organization (WHO)*^26^. This platform combines *FluNet* and *fluID* data by the *Global Influenza Surveillance and Response System (GISRS)* and national epidemiological institutions to provide world-wide yearly monitoring of influenza season severity with specification of different strains (type A: H1N1 and H3, type B: Victoria and Yamagata). The Global Influenza Programme is dependent on monitoring policies of individual countries and data is therefore not uniformly acquired over countries. After the pH1N1 many countries only implemented consistent subtyping for type A influenza infections, and type B influenza subtyping data was only available for the Netherlands in this study. Since the Global Influenza Programme reports sentinel and non-sentinel specimens that were collected after medical consultation, it also does not provide reliable estimations for mild influenza infections that generally do not require medical consultation. The Dutch National Institute for Public Health and the Environment has since 2012 annually released generalized estimates of symptomatic influenza incidence for the whole population^27,28^. These estimates provide more reliable influenza incidence rates in the whole population instead of only reporting laboratory-confirmed test results, especially for infections less frequently requiring medical attention such as type B influenza. This measure is calculated by multiplying the national influenza-like illness incidence rate with the distribution of influenza strains (derived from laboratory testing of a representative random subsample of people with influenza-like illness), while correcting for healthcare seeking behaviour.

Linear mixed models were implemented to investigate the relationships between hypersomnolence disorder onset and preceding influenza season severity. Data from the Netherlands, France, Italy and Czech Republic were first normalised per country considering the population sizes among these countries. Normalisation was performed by indexing the number of influenza infections in each year to the total infections over years in each country. Linear mixed models with country as random effect were separately performed with type A H1N1 and type A H3N2 influenza incidence rates as predictors, to predict the incidences of different CDH phenotypes. Analyses were separately performed for NT1 and the combined group of NT2/IH, and within these groups for all individuals, only children and only adults.

Additional non-parametric Spearman correlation analyses were implemented (data were not normally distributed) to respectively correlate the NT1 incidence rates with influenza season severity in individual countries. Correlational analyses were repeatedly performed for each country and for all individuals, only children and only adults. Type B Victoria and type B Yamagata data were only reliably available for the Netherlands. These analyses were not performed for the NT2/IH group because of insufficient power per country. As a sensitivity analysis to test whether our correlations could have been due to chance, we shifted incidence rates of influenza strains one year backward and repeated the correlations with respectively NT1 incidence rates in case there was a significant relationship in the main analysis. We chose a correlation coefficient > 0.2 and P-value < 0.05 as a significant correlation.

## Results

We included representative samples (Table 1) of NT1 (N = 981), NT2 (N = 189) and IH (N = 356).

**Table 1:** Population characteristics.

| Parameter | Outcome | NT1 (N=981) | NT2 (N=189) | IH (N=356) | P-value |
| --- | --- | --- | --- | --- | --- |
| Sex | Female | 474 (48.4) | 92 (49.2) | 258 (72.7) | <0.001 |
|  | Male | 505 (51.6) | 95 (50.8) | 97 (27.3) |  |
| Age | Years | 20.28 (12.14) | 21.2 (10.5) | 22.51 (11.49) | 0.005 |
| Age group # | Children | 563 (57.4) | 109 (57.7) | 171 (48.0) | 0.015 |
|  | Adults | 386 (39.3) | 80 (42.3) | 169 (47.5) |  |
| Cataplexy presence | Yes | 932 (95.3) | 0 (0) | 0 (0) | <0.001 |
|  | No | 46 (4.7) | 180 (100) | 277 (100) |  |
| Age at onset cataplexy (years) |  | 21.78 (12.48) | - | - | - |
| MSLT sleep latency (minutes) |  | 3.88 (3.53) | 5.57 (2.41) | 7.58 (3.10) | <0.001 |
| Hypocretin-1 levels (pg/mL) | <40 | 398 (74.1) | 0 (0.0) | 0 (0.0) | <0.001 |
|  | 40-110 | 110 (20.5) | 0 (0.0) | 0 (0.0) |  |
|  | 110-200 | 21 (3.9) | 10 (10.1)† | 4 (4.3)† |  |
|  | >200 | 8 (1.5) | 89 (89.9) | 88 (95.7) |  |
| HLA-DQB1*06:02 positivity | Yes | 839 (97.1) | 58 (35.2) | 35 (18.6) | <0.001 |
|  | No | 25 (2.9) | 107 (64.8) | 153 (81.4) |  |
| ESS | Score | 17.00 [14.00, 20.00] | 15.0 [12.0, 17.5] | 15.0 [12.0, 18.0] | <0.001 |

The values represent means and standard deviations in round brackets for continuous variables and percentages for categorical variables. ESS scores are presented as medians and interquartile ranges in square brackets. # 32 NT1 and 16 IH patients had missing data in their age of symptom onset. † the 14 individuals with NT2 or IH and hypocretin-1 levels between 110-200 all had hypocretin-1 levels above 150. ESS: Epworth sleepiness scale; HLA: human leukocyte antigen; IH: idiopathic hypersomnia; MSLT: multiple sleep latency test; NT1: narcolepsy type 1; NT2: narcolepsy type 2; PSG: polysomnography.

### The 2010 and 2013 NT1 incidence peaks

We identified two main incidence peaks in NT1 when including all age groups (Fig. 1A). The increased incidence was also significantly found separately for children (2.02-fold increase) and adults (1.99-fold increase) in 2010 and for children (1.84-fold increase) and adults (1.69-fold increase) in 2013 (Fig. 1B-C).

**Figure 1.**
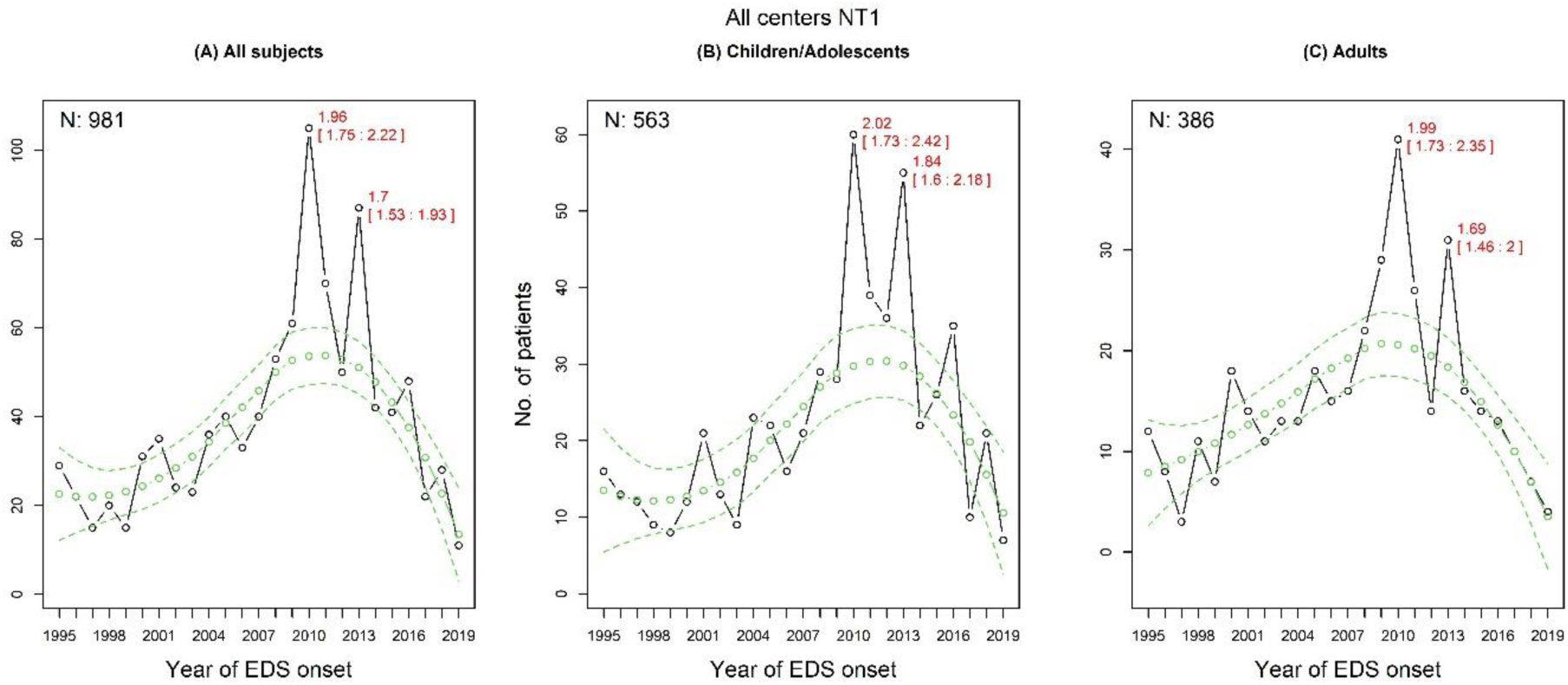
Narcolepsy type 1 incidence rates. The changes in incidence of narcolepsy type 1 (NT1) in all individuals **(A)**, children **(B)** and adults **(C)**. The predicted incidence rates and their 95% predictive confidence intervals are marked as green circles/lines and the actual values are in black circles. EDS: excessive daytime sleepiness.

Per country analyses in the largest datasets revealed that the 2010 NT1 incidence peak was significantly present in children in the Netherlands, France and Italy, and in adults in the Netherlands and France. The peak in 2013 was significantly present in children in the Netherlands, France and Italy, and in adults in the Netherlands and Italy (Supplementary material 1, Supplementary Fig. 1-4). Similar results were also found in NT1 excluding those with clinical diagnoses that had clear central disorder of hypersomnolence phenotype but did not strictly adhere to the ICSD-3 criteria (Supplementary material 2, Supplementary Fig. 7 and Fig. 9-12).

### The 2010 and 2013 NT2/IH incidence peaks

Similar significant incidence increases were found in 2010 and 2013 for NT2/IH (Fig. 2). The peak in 2010 was driven by child-onset cases compared to the 2013 peak that was driven by adults. Post-hoc exploratory analyses showed similar contributions of NT2 and IH to the 2010 and 2013 incidence peaks (Supplementary material 1, Supplementary Fig. 5-6).

**Figure 2.**
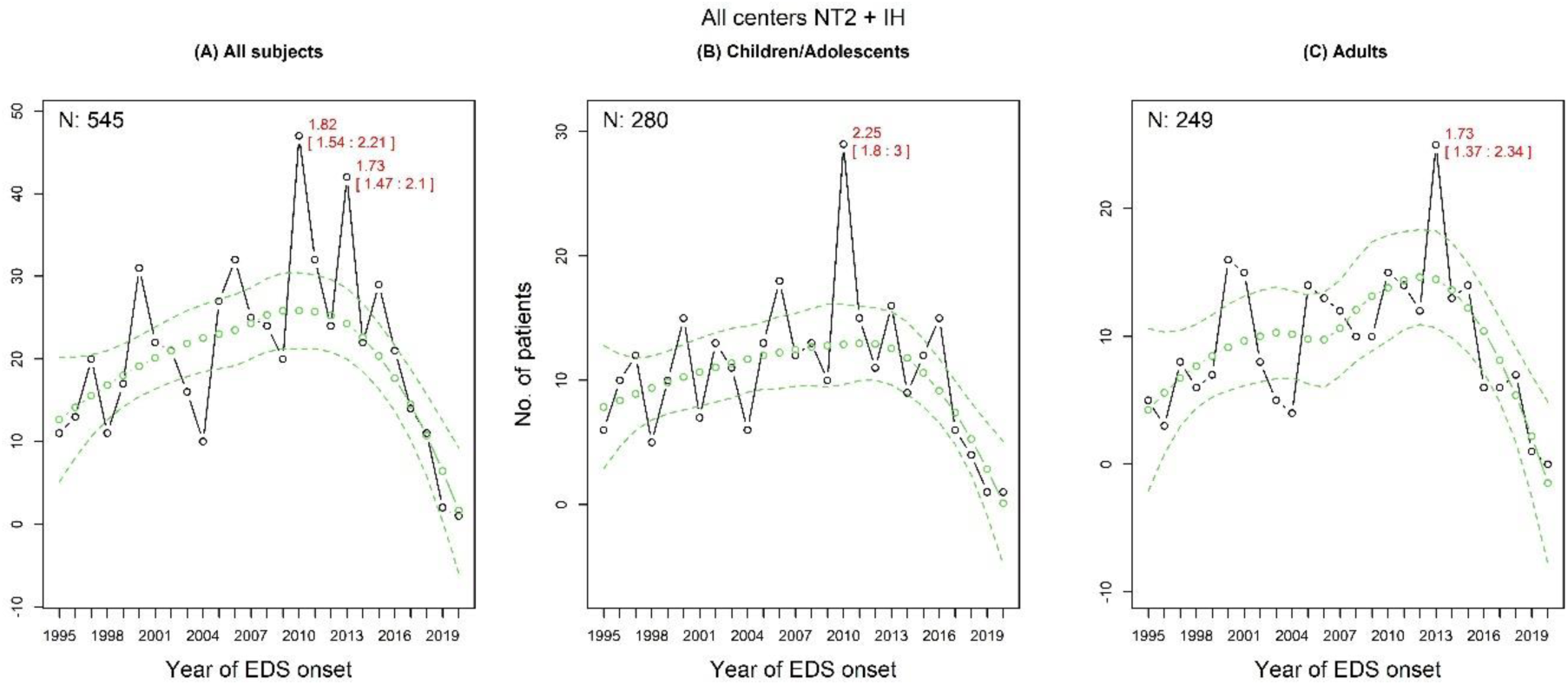
Narcolepsy type 2 and idiopathic hypersomnia incidence rates. The changes in narcolepsy type 2 (NT2) and idiopathic hypersomnia (IH) in all individuals **(A)**, children **(B)** and adults **(C)**. The predicted incidence rates and their 95% predictive confidence intervals are marked as green circles/lines and the actual values are in black circles. EDS: excessive daytime sleepiness.

The 2010 and 2013 incidence peaks in NT2/IH persisted when excluding individuals that were HLA DQB1*06:02 positive (Fig. 3). Permutation testing with 10,000 random subsamples of 90% of the children with NT2/IH resulted in consistent replication of the significant 2010 incidence peak in NT2/IH (96.4% of iterations with P-value < 0.05). The significant 2013 incidence peak in adults with NT2/IH could only be replicated in 64.1% of the iterations of the same permutation testing, suggesting this peak could be a false positive finding. Similar results were also found in NT2/IH excluding those with clinical diagnoses that had clear central disorder of hypersomnolence phenotype but did not strictly adhere to the ICSD-3 criteria (Supplementary material 2, Supplementary Fig. 8 and Fig. 13-14).

**Figure 3.**
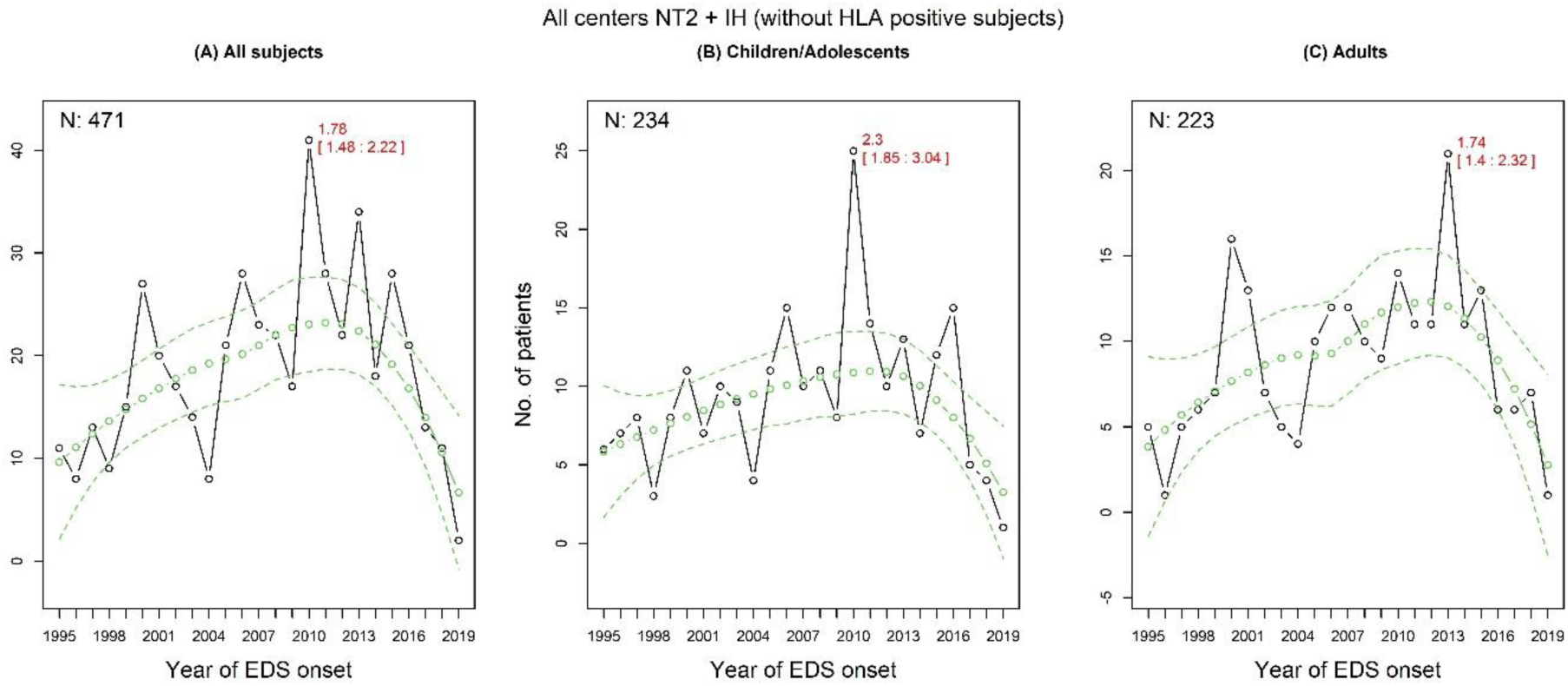
Narcolepsy type 2 and idiopathic hypersomnia incidence rates excluding HLA DQB1*06:02 positive subjects. The changes in narcolepsy type 2 (NT2) and idiopathic hypersomnia (IH) all individuals **(A)**, children **(B)** and adults **(C)**. The predicted incidence rates and their 95% predictive confidence intervals are marked as green circles/lines and the actual values are in black circles. EDS: excessive daytime sleepiness.

### NT1 incidence and preceding influenza season severity

We identified a significantly positive relationship between type A H1N1 influenza season severity and NT1 incidence in children and adults when combining data from the four largest countries in linear mixed model analysis (Table 2). A significantly negative relationship was found between type A H3N2 influenza season severity and NT1 incidence in both age groups. Similar results were also found in NT1 excluding those with clinical diagnoses that had clear central disorder of hypersomnolence phenotype but did not strictly adhere to the ICSD-3 criteria (Supplementary material 2, Supplementary Fig. 15).

**Table 2.** Results of linear mixed models.

|  | Estimate | Confidence interval (95%) | p-value |
| --- | --- | --- | --- |
| **NT1 and type A H1N1** |  |  |  |
| All subjects | 0.0017 | [0.0011, 0.0022] | <.0001 |
| Children | 0.0008 | [0.0004, 0.0013] | 0.0001 |
| Adults | 0.0008 | [0.0006, 0.001] | <.0001 |
| **NT1 and type A H3N2** |  |  |  |
| All subjects | -0.0034 | [-0.006, -0.0008] | 0.005 |
| Children | -0.002 | [-0.004, -0.0003] | 0.011 |
| Adults | -0.0014 | [-0.0024, -0.0001] | 0.009 |
| **NT2+IH and type A H1N1** |  |  |  |
| All subjects | 0.0005 | [0.0002, 0.0008] | 0.0007 |
| Children | 0.0004 | [0.0002, 0.0005] | <0.0001 |
| Adults | 0.0002 | [0.0001, 0.0004] | 0.0738 |
| **NT2+IH and type A H3N2** |  |  |  |
| All subjects | -0.0009 | [-0.0019, 0.0004] | 0.1308 |
| Children | -0.0007 | [-0.0014, -0.00006] | 0.0341 |
| Adults | -0.0002 | [-0.0007, 0.0009] | 0.9567 |

IH: idiopathic hypersomnia; NT1: narcolepsy type 1; NT2: narcolepsy type 2.

Per country correlational analyses revealed that the type A influenza effects were strongest in the Netherlands and France. In the Netherlands we additionally identified a strong positive correlation with type B Victoria influenza severity in children (correlation coefficient 0.94, P-value < 0.001, as shown in Fig. 4). When shifting incidence rates of influenza strains one year backward as a sensitivity analysis, no significant correlations remained present between type A H1N1, type A H3N2 or type B Victoria, and NT1 incidence rates (Supplementary material 2, Supplementary Fig. 16).

**Figure 4.**
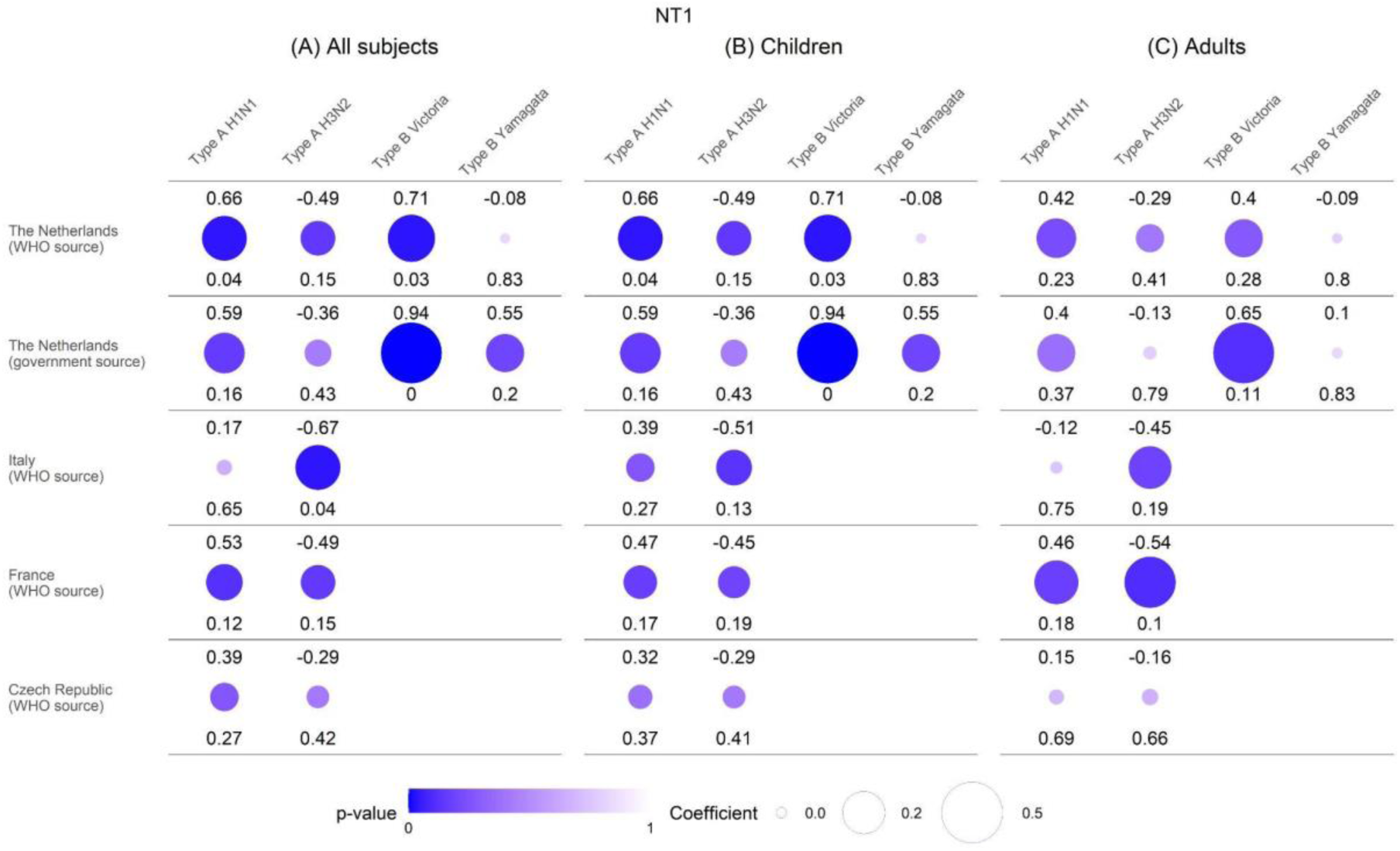
Narcolepsy type 1 and preceding influenza season severity correlations per country. The three panels respectively represent the correlations between annual incidence rates of each influenza strain incidence and narcolepsy type 1 (NT1) incidence in the following year for all individuals **(A)**, children **(B)** and adults **(C)**. Top values and circle sizes represent the correlation coefficients. Bottom values and circle colours represent the level of significance of the correlation.

### NT2/IH incidence and preceding influenza season severity

A significantly positive relationship was seen for type A H1N1 influenza severity and paediatric NT2 and IH incidence when combining data from all countries (Table 2). A similar significantly negative correlation was seen for type A H3N2 and children with NT2/IH. Significant associations between influenza season severity and NT2/IH incidence were not present for adults. Similar results were found when excluding NT2/IH with a clinical diagnosis.

## Discussion

We replicated the 2010 and 2013 NT1 incidence peaks in complete datasets from multiple European countries. Both peaks in 2010 and 2013 were reported in children and adults. Similar incidence peaks were seen for NT2 and IH in 2010 with a smaller peak in 2013. These peaks were driven by children in 2010 and by adults in 2013. Regression analyses showed that the NT1 incidence in children had a significantly positive association with type A H1N1 influenza season severity but a negative association with type A H3N2 influenza season severity. This effect was most pronounced in the Netherlands, where influenza type B Victoria was even more strongly correlated with NT1 incidence in children. Paediatric NT2 and IH incidence was significantly positively related to preceding type A H1N1 influenza season and negatively related to type A H3N2 influenza severity.

Besides the significant relationship with type A H1N1 influenza, we identified a strong correlation between NT1 and influenza type B Victoria incidence rates. It is plausible that other infections have been responsible for triggering NT1 before the reintroduction of the type A H1N1 influenza virus in 2009. Streptococcal infections have previously been proposed but supporting evidence remains inconclusive^11–15^. Other flu strains are likely candidates for triggering NT1 and we here provide first epidemiological evidence that infection with influenza type B Victoria could be an important new suspect^18,29^. Influenza type B Victoria has consistently circulated in the past century and is known to more frequently and heavily affect children compared to adults^30,31^. This matches the typical adolescent onset of NT1^32^. The 2013 NT1 incidence peak is more pronounced in children compared to the peak in 2010. When reviewing preceding flu season severity and influenza strain dominance, it is a possibility that the peak in 2010 was more frequently triggered by the type A H1N1 influenza virus, and the 2013 peak mainly by influenza type B Victoria. External testing of this hypothesis is possible when comparing recently published Chinese NT1 annual onset rates with preceding Chinese flu season severity from the WHO Global Influenza Programme^21,26^. In China, there were relative NT1 child-onset incidence peaks in 2010, 2012 and 2014 and type B Victoria was notably among the dominant influenza strains in all three preceding flu seasons (compared to type A H1N1 that was not dominant in the 2011-2012 influenza season). Similar comparisons could not be performed for the United States where type B influenza is generally not subtyped^26^. Growing bodies of evidence indicate links between influenza infection and the onset of various other neurological disorders with a typical later disease onset, including Parkinson’s Disease, dementia, and multiple sclerosis^33^. Our findings fit the hypothesis that there is a limited pool of genetically susceptible individuals (that are HLA-DQB1*06:02 positive) who might develop NT1 after an immunological trigger (such as different flu strains, streptococcal infections, and Pandemrix vaccination). After an NT1 incidence peak, this pool of remaining susceptible individuals shrinks, and the NT1 incidence rate in the following year drops. A nuanced perspective is essential in approaching these hypotheses and further research is needed to establish any causal connections between NT1, immunological triggers, and type B Victoria influenza in particular.

In our study, we observed a notably adverse relationship between NT1 incidence and preceding H3N2 influenza season severity. Influenza strains are recognized for their competitive circulation in each influenza season, a process mediated by the speed of genetic recombination of individual strains, the lingering antibody titers from the last circulation of a specific influenza strain. Partial or cross-immunity between influenza type A (H1N1 and H3N2) and type B (Victoria and Yamagata) also contributes to these effects^34–36^. This interplay of factors typically results in the dominance of one or two influenza strains during each season^37^. Within this competitive environment it seems logical that there is less space for influenza strains that could trigger NT1 when non-triggering influenza strains are dominant in a particular year. Therefore, we hypothesize that the negative correlation with H3N2 influenza season severity could be a protective phenomenon for other (possibly NT1-inducing) influenza strains.

NT1 incidence fluctuations so far seemed more pronounced in children. Using complete incidence data from seven European countries we have now also uncovered that there has been a significant peak in adult-onset NT1 in 2010 following the pH1N1. This suggests that Pandemrix vaccination and/or type A H1N1 influenza infection could also trigger NT1 in genetically susceptible adults. Similar findings have been reported by Dauvilliers et al.^4^. The 2010 adult-onset NT1 peak was not identified in most earlier studies which could be related to the relatively smaller magnitude of the 2010-peak in adults, and the increased awareness that primarily focused on child-onset NT1 following the pH1N1^38,39^. Pandemrix was also mainly used in younger age groups and a relationship with type A H1N1 influenza infection was only suggested later^39,40^.

We are the first to identify incidence fluctuations in NT2 and IH with peaks in 2010 in children and in 2013 in adults. The 2010 peak in children remained present in our sensitivity analyses (excluding HLA DQB1*06:02 positive individuals and during permutation test), suggesting this peak is not due to misdiagnosis of people with NT1 or dependent on statistical methods. Our results suggest an influence of annually fluctuating external factors similar to NT1. Using the same sample from the Netherlands, we have recently uncovered that individuals with NT2 and IH also regularly report respiratory infections close to onset of their excessive daytime sleepiness^23^. Together with the significantly positive relationship with preceding type A H1N1 influenza season severity, this suggests that a trigger-related immunological basis underlying NT2 and IH pathophysiology should be seriously considered, and warrants future investigations^23^. Media attention for EDS after Pandemrix-associated onset of NT1 could have played a role in the 2010 peak in NT2 and IH with more individuals subsequently seeking medical attention^38,39^. The absence of similar media coverage in 2013 suggests increased awareness alone cannot fully explain the peaks in NT2 and IH during that time. In our study, we cannot dismiss the possibility of influenza vaccination, especially Pandemrix, playing a delayed role in the incidence peaks. However, the link between the severity of the influenza season and the occurrence of central disorders of hypersomnolence also implies that influenza vaccination (non-Pandemrix) might offer protection against the development of hypersomnolence disorders.

There are several limitations to our study. First, the observed effects in our study were strongest in the analyses combining data from all countries and similar results were not always observed in each individual country. We believe this is most likely because of the limited data points per country (often only nine years) and the large variability in influenza monitoring policies per country^41^. Many countries improved their influenza surveillance system during pH1N1 and since then mainly focused on subtyping of type A influenza strains^42^. The Netherlands is known for its precise governmental influenza monitoring program with representative national coverage and standard subtyping of all four influenza strains through a combination of sentinel, non-sentinel and modelling-based estimates of influenza infections^27,28^. Together with the compactness of the Netherlands resulting in limited provincial influenza strain circulation differences, we believe that this likely explains the large effect sizes that we observed within the Netherlands. The immunological mechanisms responsible for the connections we identified between the onset of hypersomnolence disorder and type A H1N1, and NT1 onset with type B Victoria, remain unclear. We provide new epidemiological associations and this should not be mistaken for causality. Various underlying pathophysiological mechanisms have been proposed. While molecular mimicry may initiate a self-reactive T cell response related to hypocretin after influenza infection or Pandemrix vaccination, current supporting evidence is inconsistent. Autoaggressive bystander activation by T cells without direct cross-reactivity has also been proposed and warrants further studies^10,43^. Immunological investigations are essential to better comprehend these processes and establish possible causal relationships between infuenza infection and onset of central disorders of hypersomnolence. In such studies it is important to integrate other pathogens (such as streptococcus pyogenes and influenza type C) and different clades/sub-clades of type A and type B influenza. Our sample exhibits a minor overlap of approximately 25% with the EU-NN database, thus it cannot be regarded as entirely independent. Within this study we were also unable to split NT2 and IH for the correlational analyses with preceding influenza season severity because of limited sample sizes. Given that both diagnoses contributed comparably to the observed 2010 and 2013 incidence peaks, this could suggest a similar contribution to the observed type A H1N1 and H3N2 influenza severity relationships. For individuals with NT2 and IH, it might have been more challenging to accurately identify the onset of EDS, which could have potentially influenced our findings.

Our study unveils compelling new insights into the epidemiological dynamics of NT1, NT2, and IH across European countries, highlighting distinct incidence peaks in 2010 and 2013. Both hypersomnolence groups exhibited a significantly positive relationship with preceding type A H1N1 influenza season severity, with NT1 additionally associated with influenza type B Victoria. We propose influenza type B Victoria as a potential new candidate for triggering NT1, especially considering its frequent and relatively severe impact on children. Our results also provide clues that suggest an immune-related aetiology of NT2 and IH, and hereby offer highly anticipated insights for future investigations aiming to unravel the pathophysiologies of these enigmatic sleep disorders.

## Data Availability

All data produced in the present study are available upon reasonable request to the authors

## Supplementary material 1

### Narcolepsy type 1 incidence rates per country

**Supplementary figure 1.**
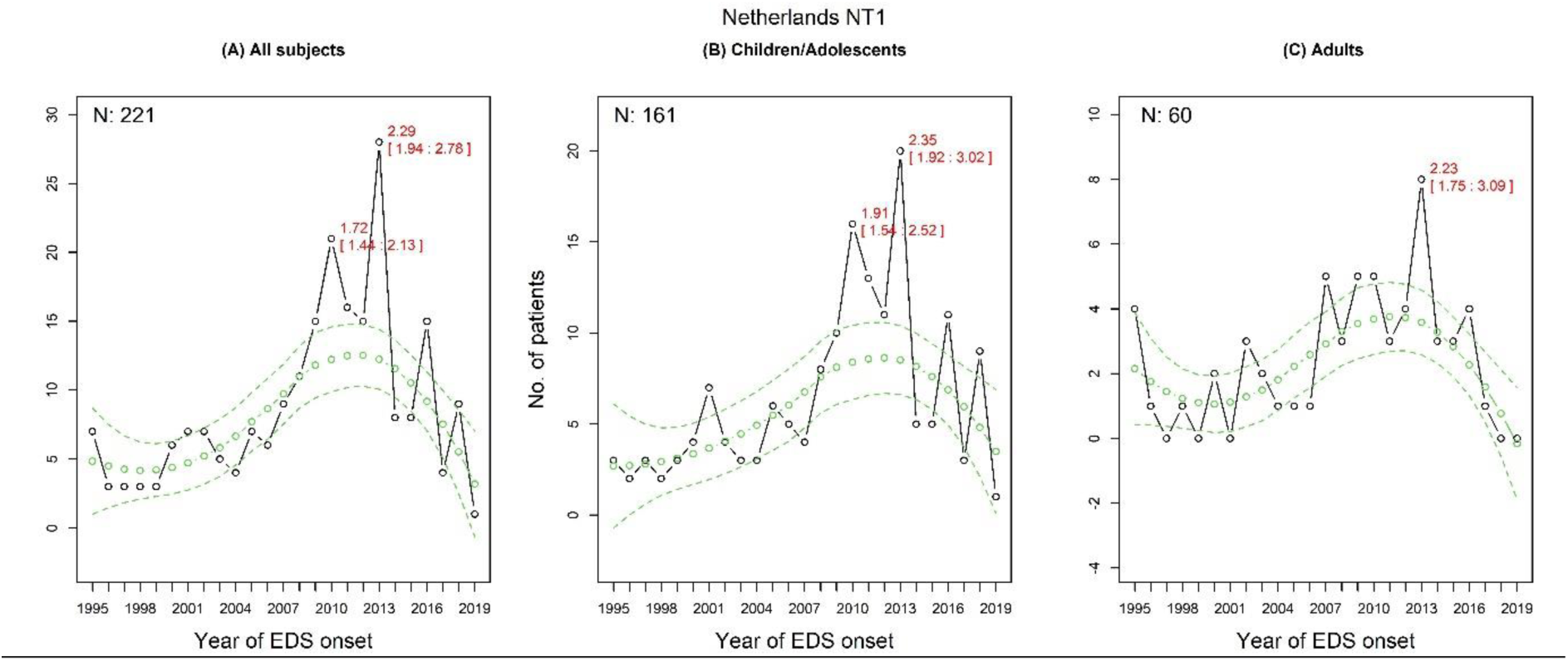
Narcolepsy type 1 incidence rates in the Netherlands. The changes in incidence of narcolepsy type 1 (NT1) in all individuals **(A)**, children **(B)** and adults **(C)**. The predicted incidence rates and their 95% predictive confidence intervals are marked as green circles/lines and the actual values are in black circles. EDS: excessive daytime sleepiness.

**Supplementary figure 2.**
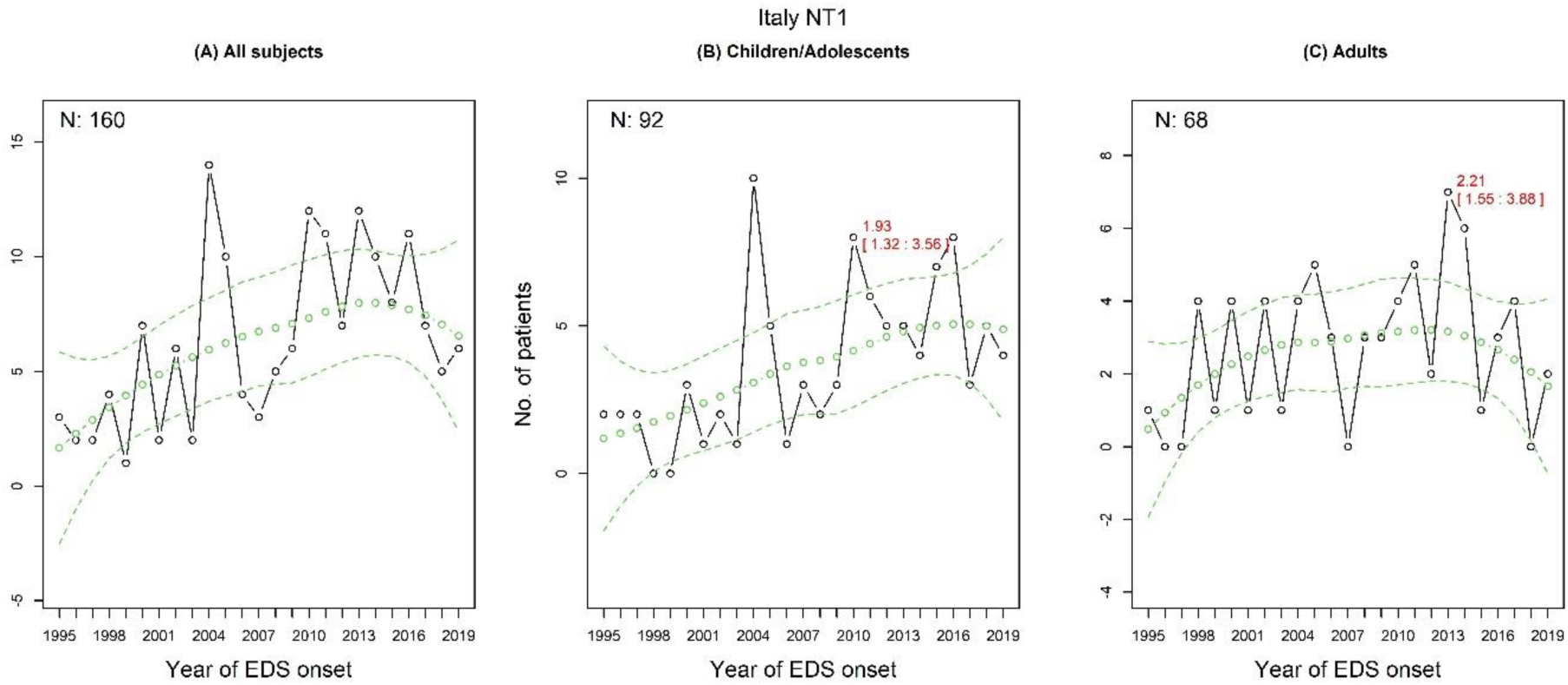
Narcolepsy type 1 incidence rates in Italy. The changes in incidence of narcolepsy type 1 (NT1) in all individuals **(A)**, children **(B)** and adults **(C)**. The predicted incidence rates and their 95% predictive confidence intervals are marked as green circles/lines and the actual values are in black circles. EDS: excessive daytime sleepiness.

**Supplementary figure 3.**
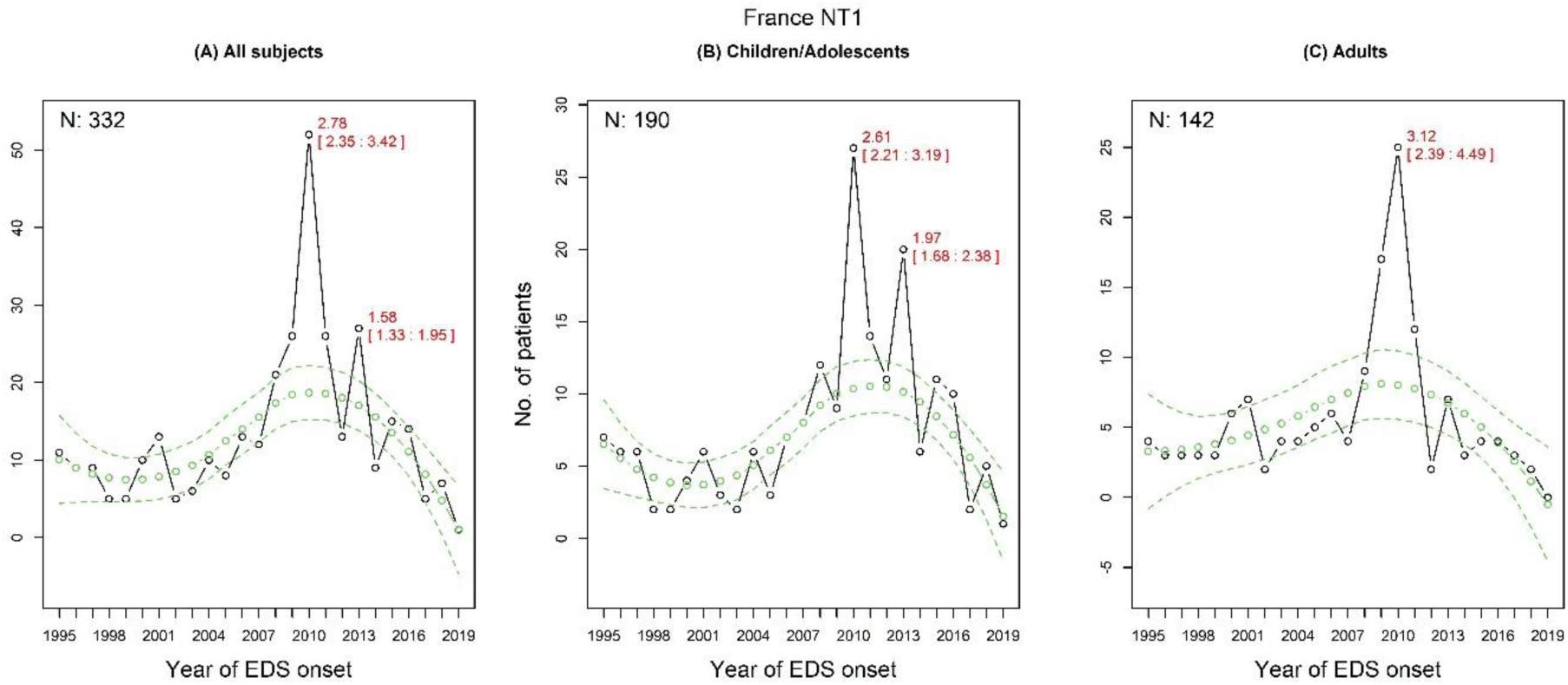
Narcolepsy type 1 incidence rates in France. The changes in incidence of narcolepsy type 1 (NT1) in all individuals **(A)**, children **(B)** and adults **(C)**. The predicted incidence rates and their 95% predictive confidence intervals are marked as green circles/lines and the actual values are in black circles. EDS: excessive daytime sleepiness.

**Supplementary figure 4.**
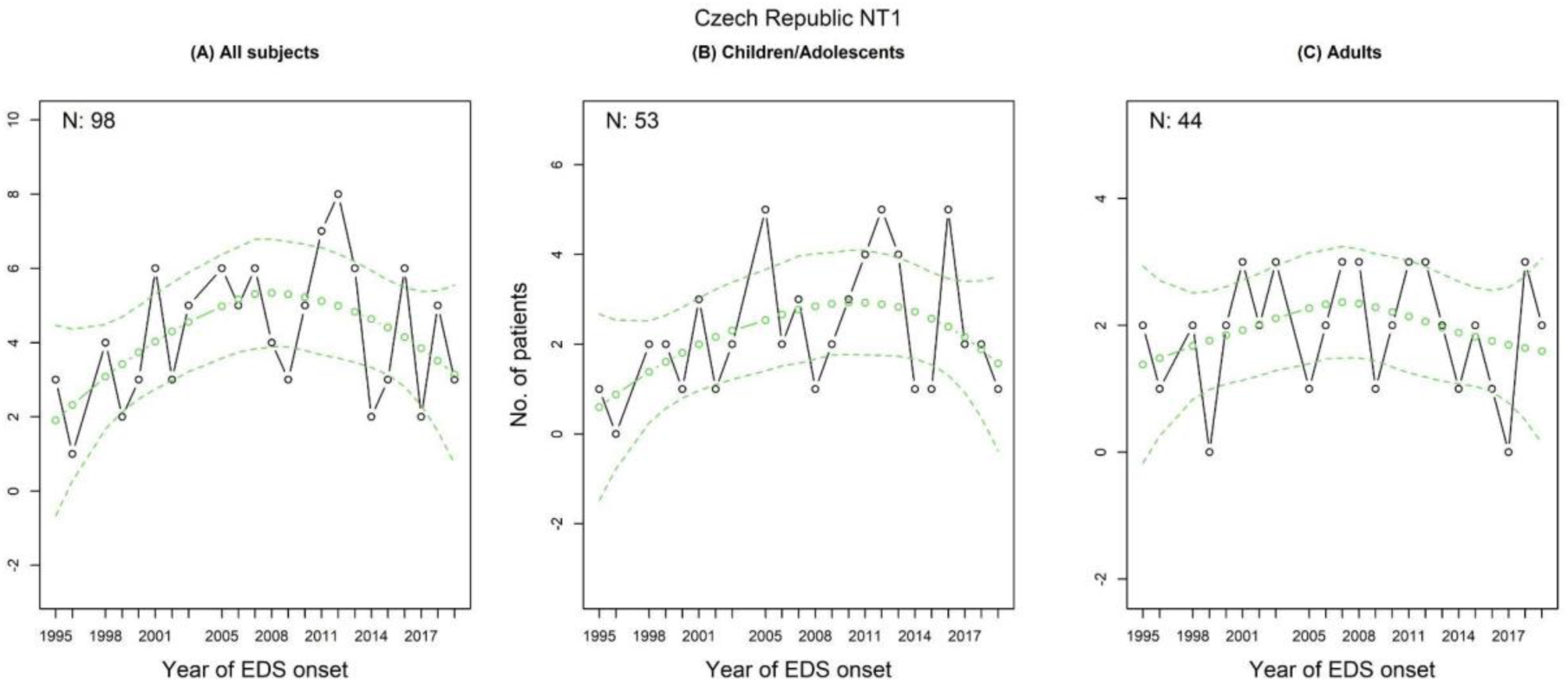
Narcolepsy type 1 incidence rates in Czech Republic. The changes in incidence of narcolepsy type 1 (NT1) in all individuals **(A)**, children **(B)** and adults **(C)**. The predicted incidence rates and their 95% predictive confidence intervals are marked as green circles/lines and the actual values are in black circles. EDS: excessive daytime sleepiness.

### Split incidence rates for narcolepsy type 2 and idiopathic hypersomnia

**Supplementary figure 5.**
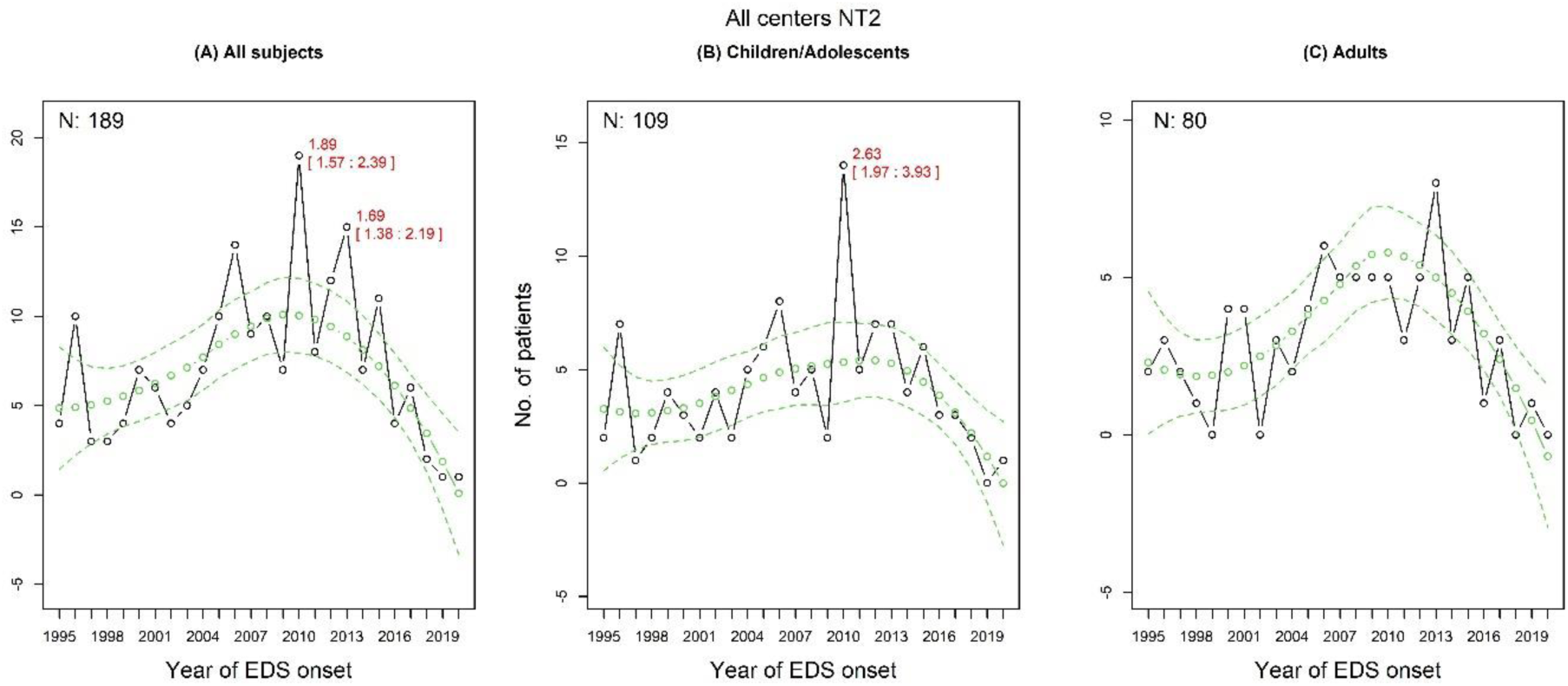
Narcolepsy type 2 incidence rates. The changes in incidence of narcolepsy type 2 (NT2) in all individuals **(A)**, children **(B)** and adults **(C)**. The predicted incidence rates and their 95% predictive confidence intervals are marked as green circles/lines and the actual values are in black circles. EDS: excessive daytime sleepiness.

**Supplementary figure 6.**
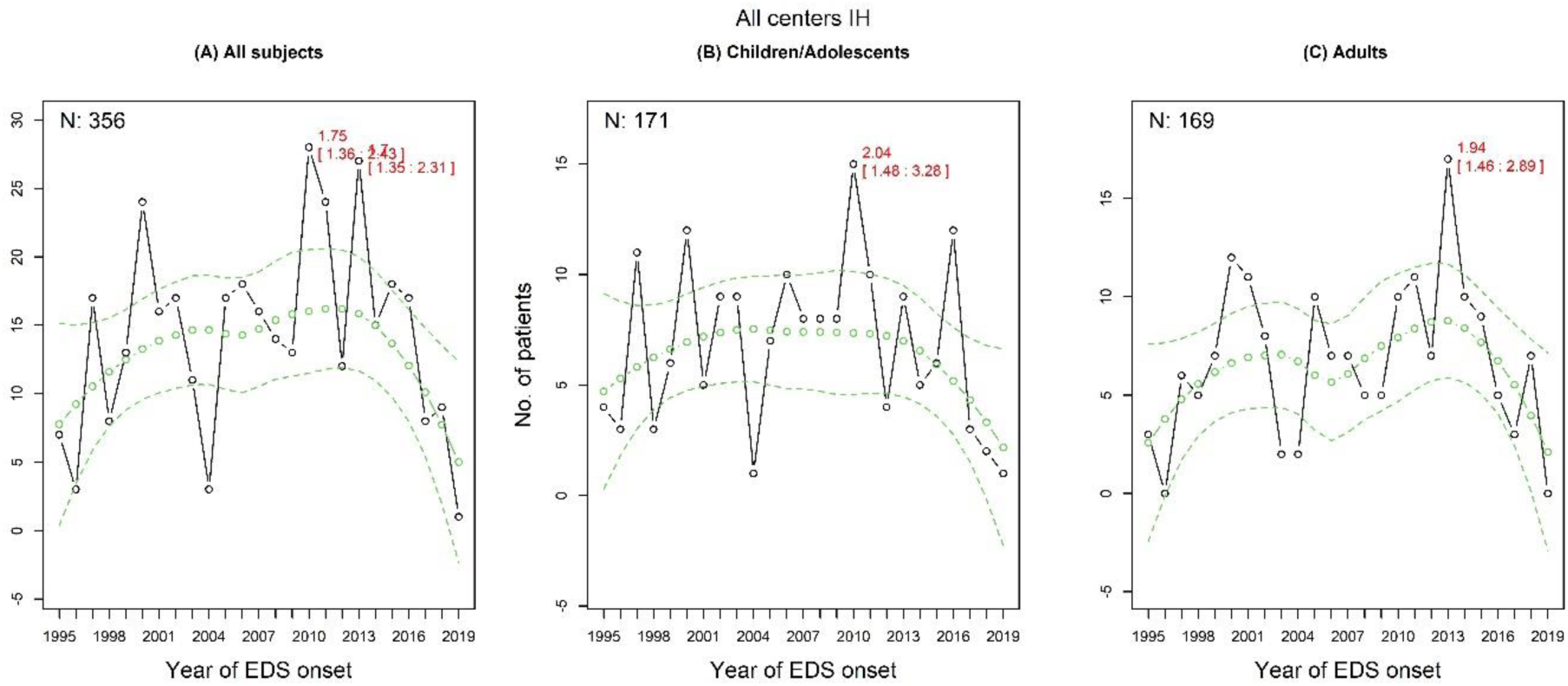
Idiopathic hypersomnia incidence rates. The changes in incidence of idiopathic hypersomnia (IH) in all individuals **(A)**, children **(B)** and adults **(C)**. The predicted incidence rates and their 95% predictive confidence intervals are marked as green circles/lines and the actual values are in black circles. EDS: excessive daytime sleepiness.

## Supplementary material 2

### Clinical diagnosis definitions

Clinical diagnosis of NT1: people with a hypocretin-1 level between 110 and 150, or people with typical cataplexy and a multiple sleep latency test sleep latency between eight and 12 minutes or just one sleep-onset rapid eye movement (REM) period.

Clinical diagnosis of NT2: people without cataplexy, at least two sleep-onset REM periods and a sleep latency between eight and 12 minutes.

Clinical diagnosis of IH: people without cataplexy, fewer than two sleep-onset REM periods and a sleep latency between eight and 12 minutes.

### Number of subjects per center

| Location | Narcolepsy type 1 |  |  | Narcolepsy type 2 |  |  | Idiopathic hypersomnia |  |  |
| --- | --- | --- | --- | --- | --- | --- | --- | --- | --- |
|  | All | Children | Adults | All | Children | Adults | All | Children | Adults |
| Heemstede, the Netherlands | 221 | 161 | 60 | 23 | 12 | 11 | 51 | 29 | 22 |
| Bologna, Italy | 160 | 92 | 68 | 25 | 14 | 11 | 61 | 21 | 40 |
| Montpellier, France | 332 | 190 | 142 | 109 | 67 | 42 | 178 | 103 | 75 |
| Prague, Czech Republic | 98 | 53 | 44 | 27 | 14 | 13 | 31 | 13 | 18 |
| Warsaw, Poland | 94 | 46 | 48 | - | - | - | - | - | - |
| Innsbruck, Austria | 29 | - | - | - | - | - | - | - | - |
| Kosice, Slovak Republic | 27 | 11 | 14 | 5 | 2 | 3 | 5 | 1 | 4 |
| Madrid, Spain | 20 | 10 | 10 | - | - | - | 14 | 4 | 10 |

### Results excluding those with a clinical diagnosis

Comparable outcomes were observed when excluding individuals with a clinical diagnosis in NT1 (Supplementary material 2, Supplementary Fig. 7). The 2010 incidence peak in NT2/IH persisted in children when excluding individuals with a clinical diagnosis (Supplementary material 2, Supplementary Fig. 8). In each country, similar results were also seen when excluding individuals with a clinical diagnosis (Supplementary material 2, Supplementary Fig. 9-12).

**Supplementary figure 7.**
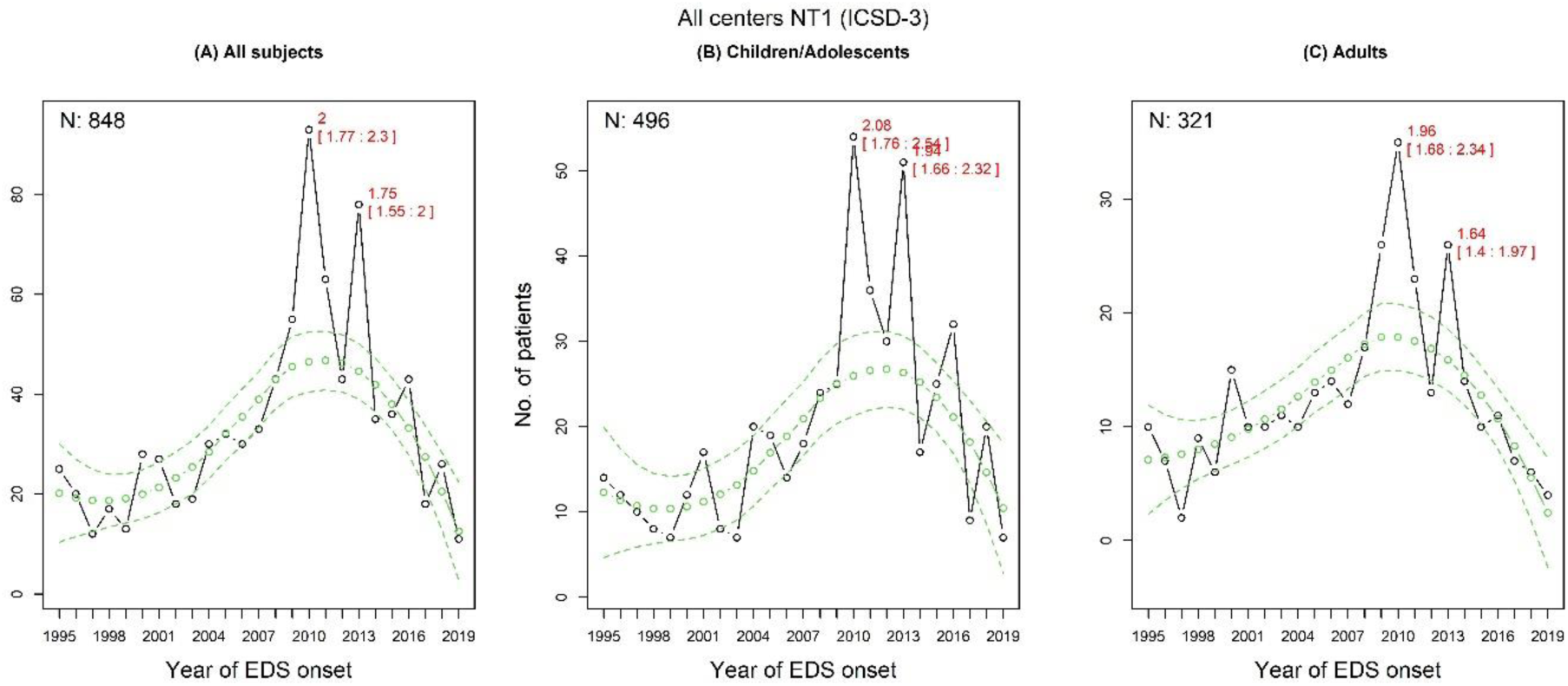
Narcolepsy type 1 incidence rates excluding clinical diagnosis. The changes in narcolepsy type 1 (NT1) in all individuals **(A)**, children **(B)** and adults **(C)**. The predicted incidence rates and their 95% predictive confidence intervals are marked as green circles/lines and the actual values are in black circles. EDS: excessive daytime sleepiness.

**Supplementary figure 8.**
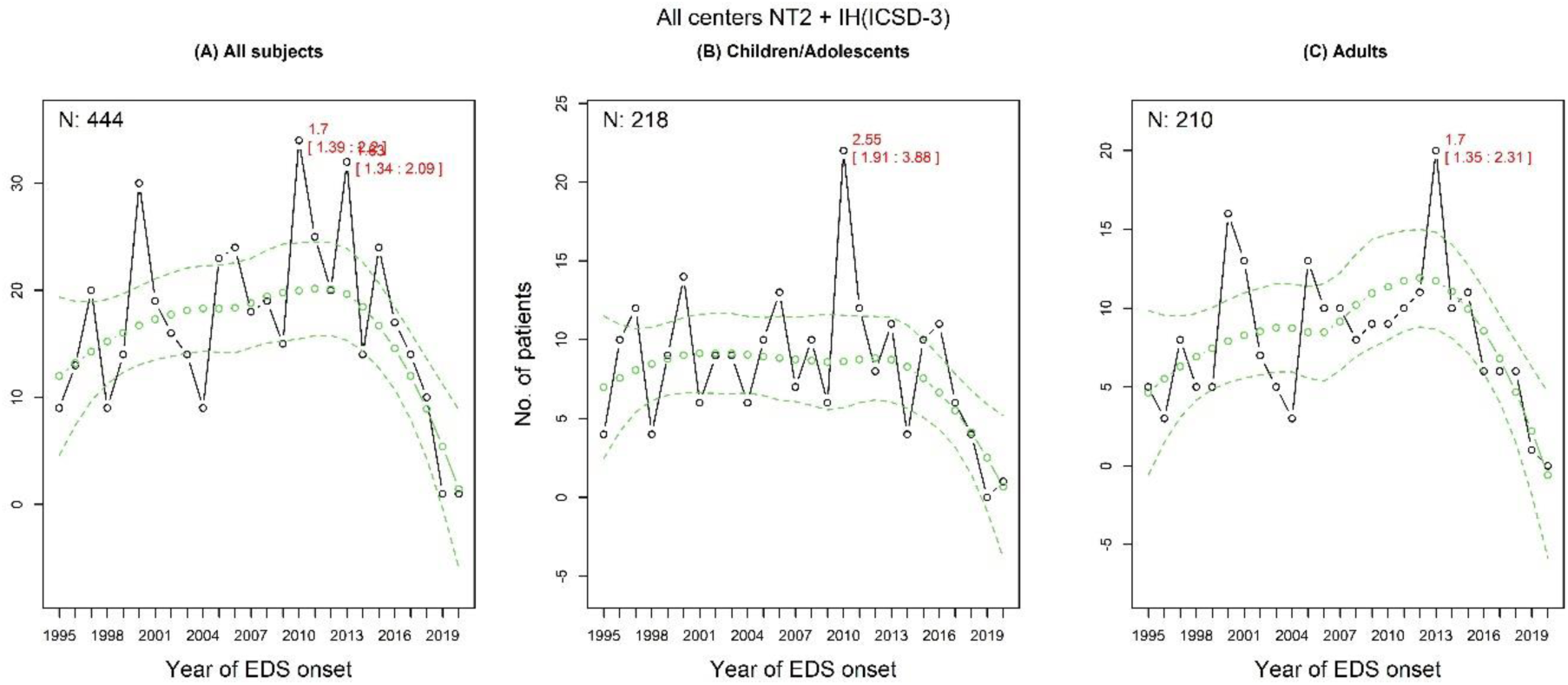
Narcolepsy type 2 and idiopathic hypersomnia incidence rates excluding clinical diagnosis. The changes in narcolepsy type 2 (NT2) and idiopathic hypersomnia (IH) in all individuals **(A)**, children **(B)** and adults **(C)**. The predicted incidence rates and their 95% predictive confidence intervals are marked as green circles/lines and the actual values are in black circles. EDS: excessive daytime sleepiness.

### Narcolepsy type 1 incidence rates per country excluding clinical diagnosis

**Supplementary figure 9.**
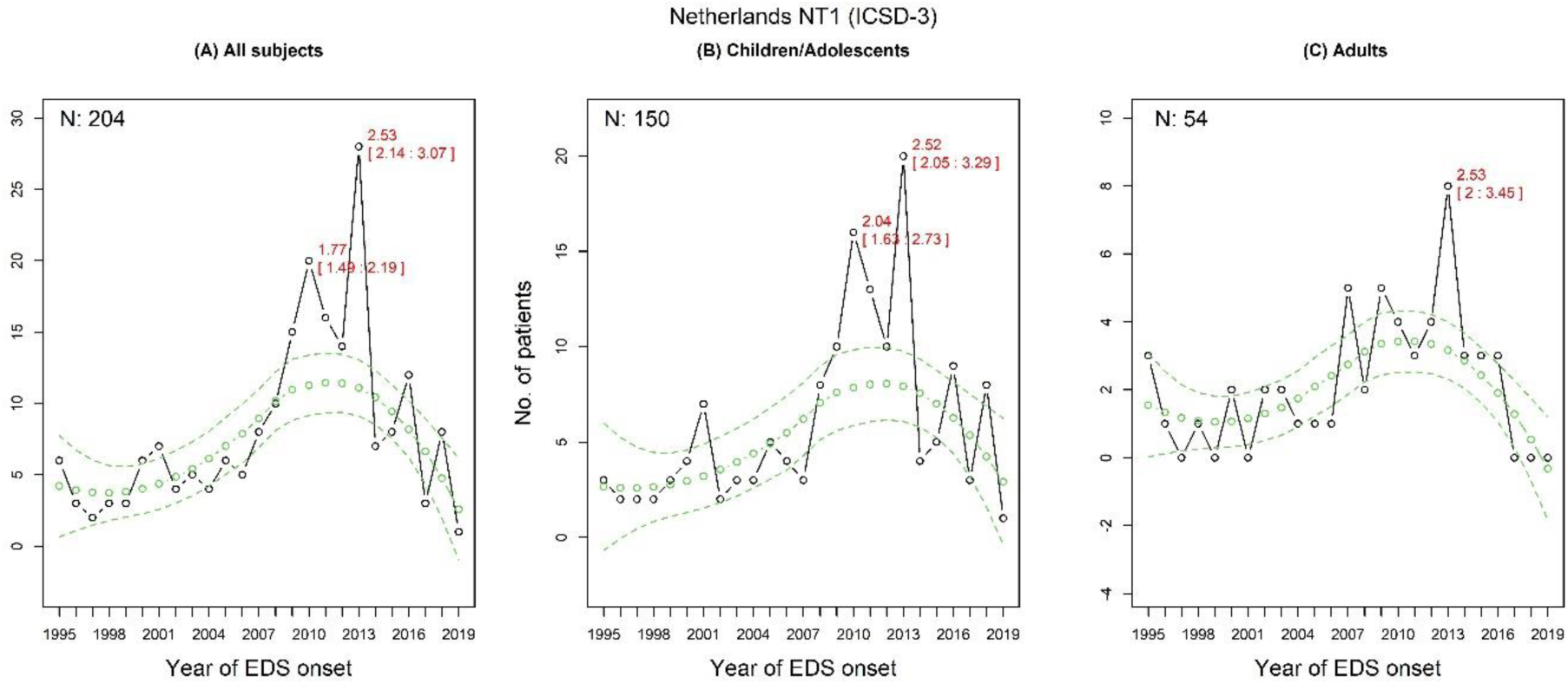
Narcolepsy type 1 incidence rates in the Netherlands excluding clinical diagnosis. The changes in narcolepsy type 1 (NT1) in all individuals **(A)**, children **(B)** and adults **(C)**. The predicted incidence rates and their 95% predictive confidence intervals are marked as green circles/lines and the actual values are in black circles. EDS: excessive daytime sleepiness.

**Supplementary figure 10.**
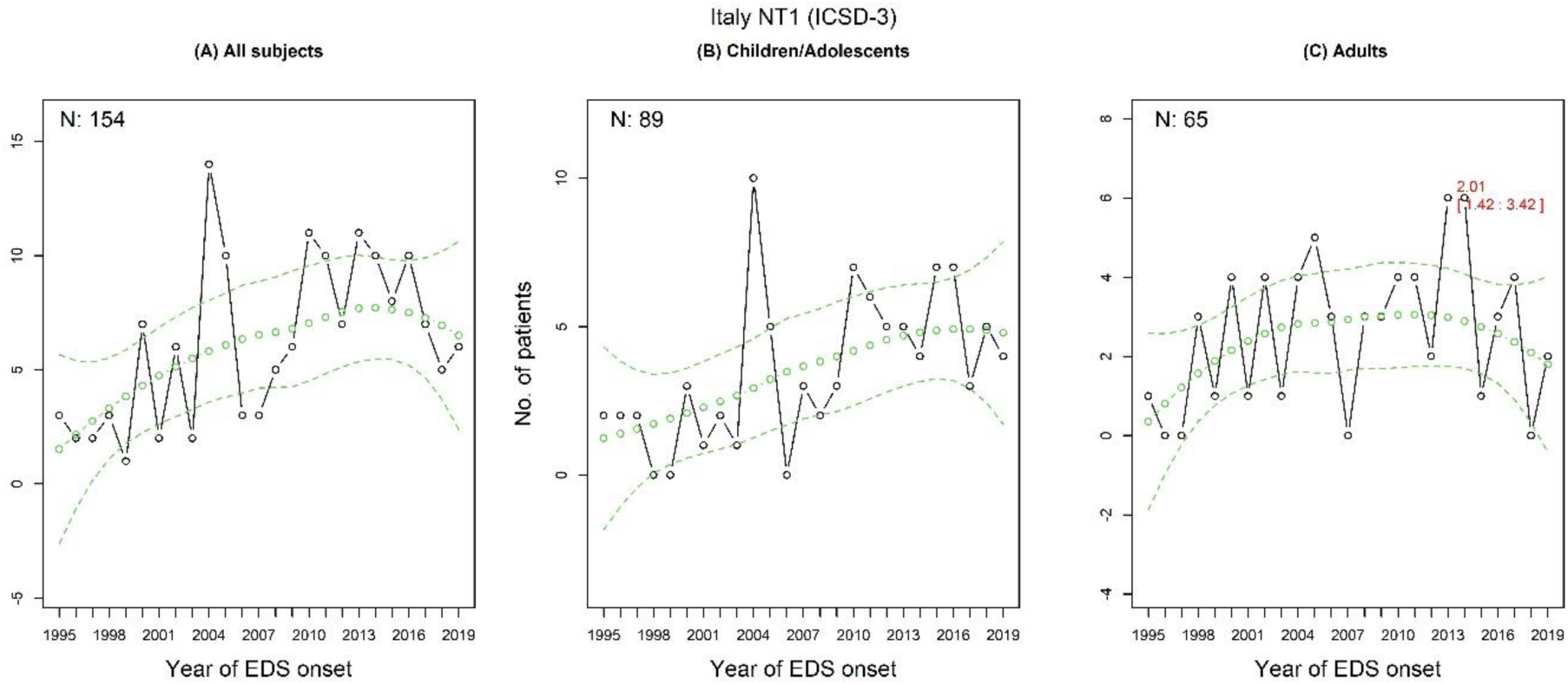
Narcolepsy type 1 incidence rates in Italy excluding clinical diagnosis. The changes in narcolepsy type 1 (NT1) in all individuals **(A)**, children **(B)** and adults **(C)**. The predicted incidence rates and their 95% predictive confidence intervals are marked as green circles/lines and the actual values are in black circles. EDS: excessive daytime sleepiness.

**Supplementary figure 11.**
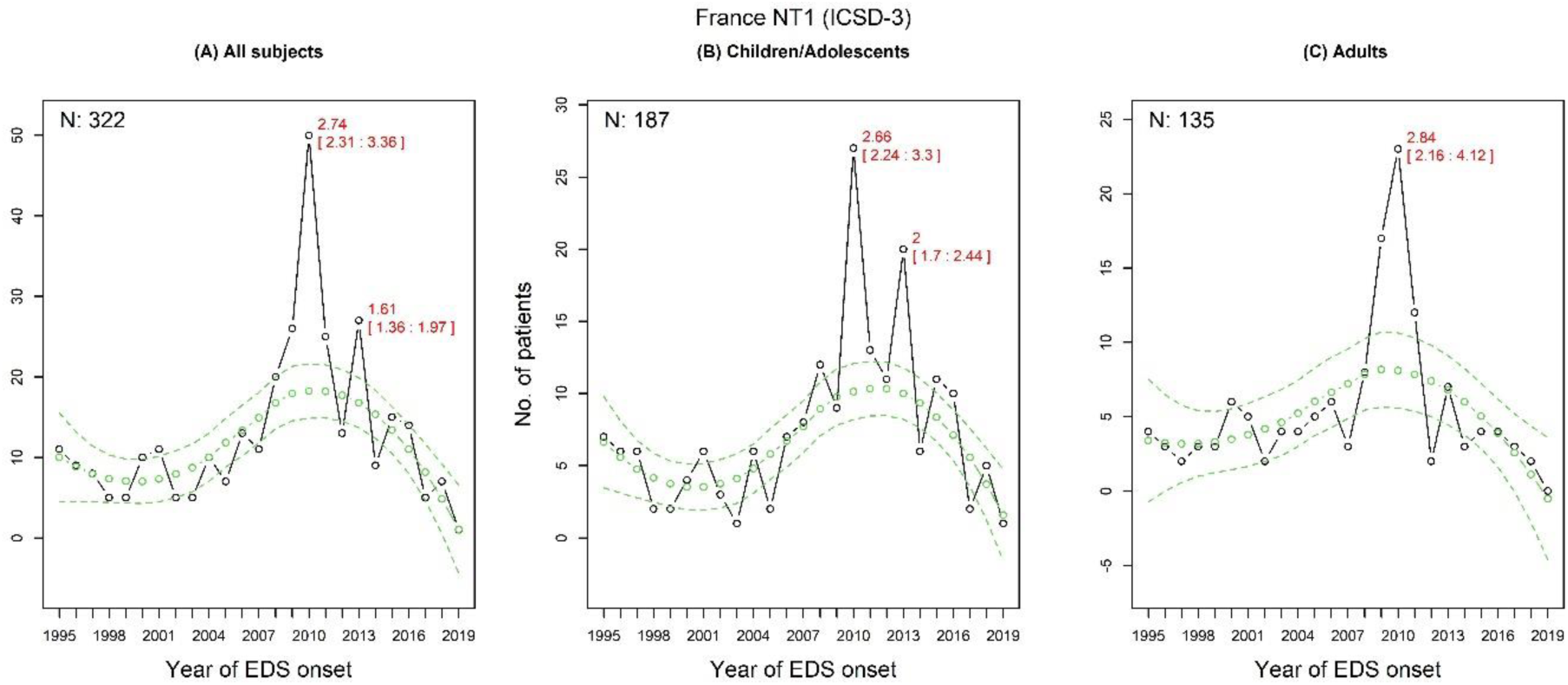
Narcolepsy type 1 incidence rates in France excluding clinical diagnosis. The changes in narcolepsy type 1 (NT1) in all individuals **(A)**, children **(B)** and adults **(C)**. The predicted incidence rates and their 95% predictive confidence intervals are marked as green circles/lines and the actual values are in black circles. EDS: excessive daytime sleepiness.

**Supplementary figure 12.**
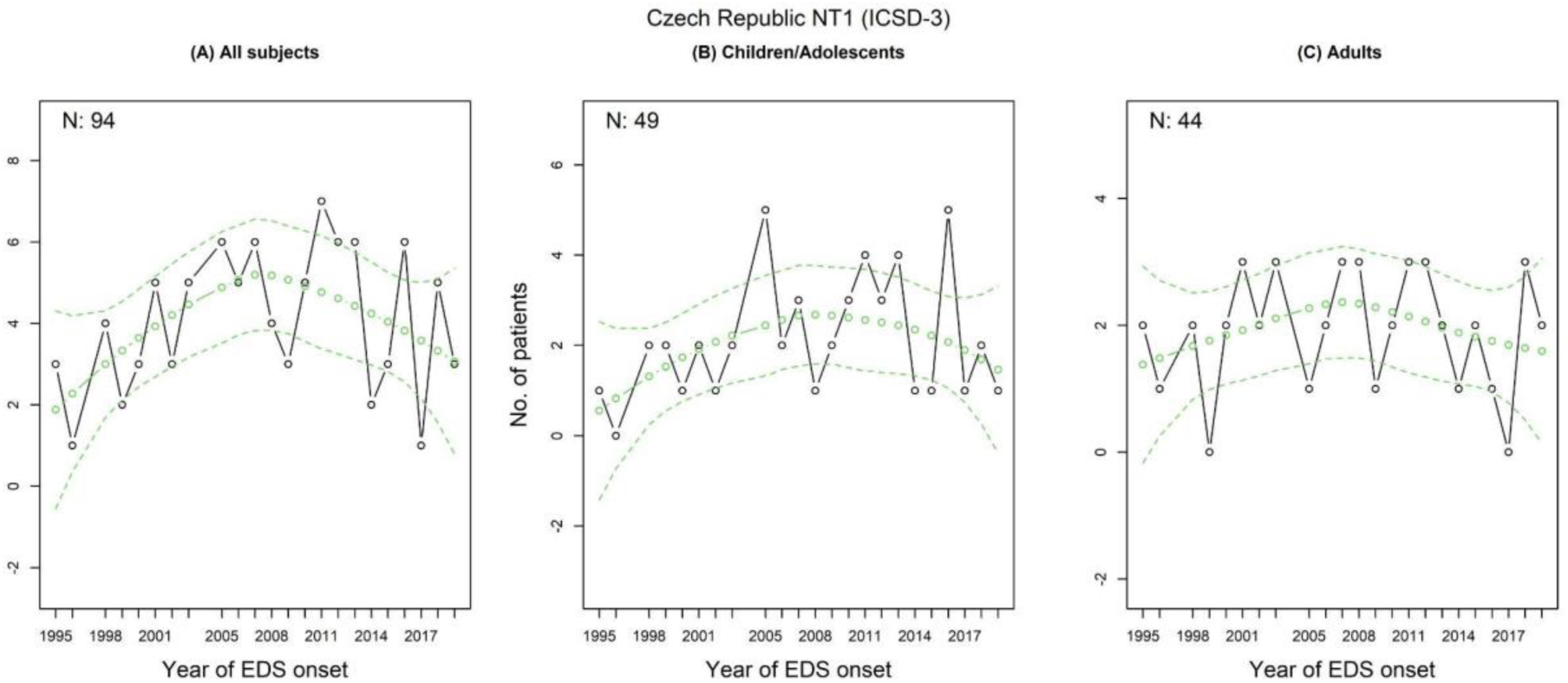
Narcolepsy type 1 incidence rates in Czech Republic excluding clinical diagnosis. The changes in narcolepsy type 1 (NT1) in all individuals **(A)**, children **(B)** and adults **(C)**. The predicted incidence rates and their 95% predictive confidence intervals are marked as green circles/lines and the actual values are in black circles. EDS: excessive daytime sleepiness.

### Split incidence rates for narcolepsy type 2 and idiopathic hypersomnia excluding clinical diagnosis

**Supplementary figure 13.**
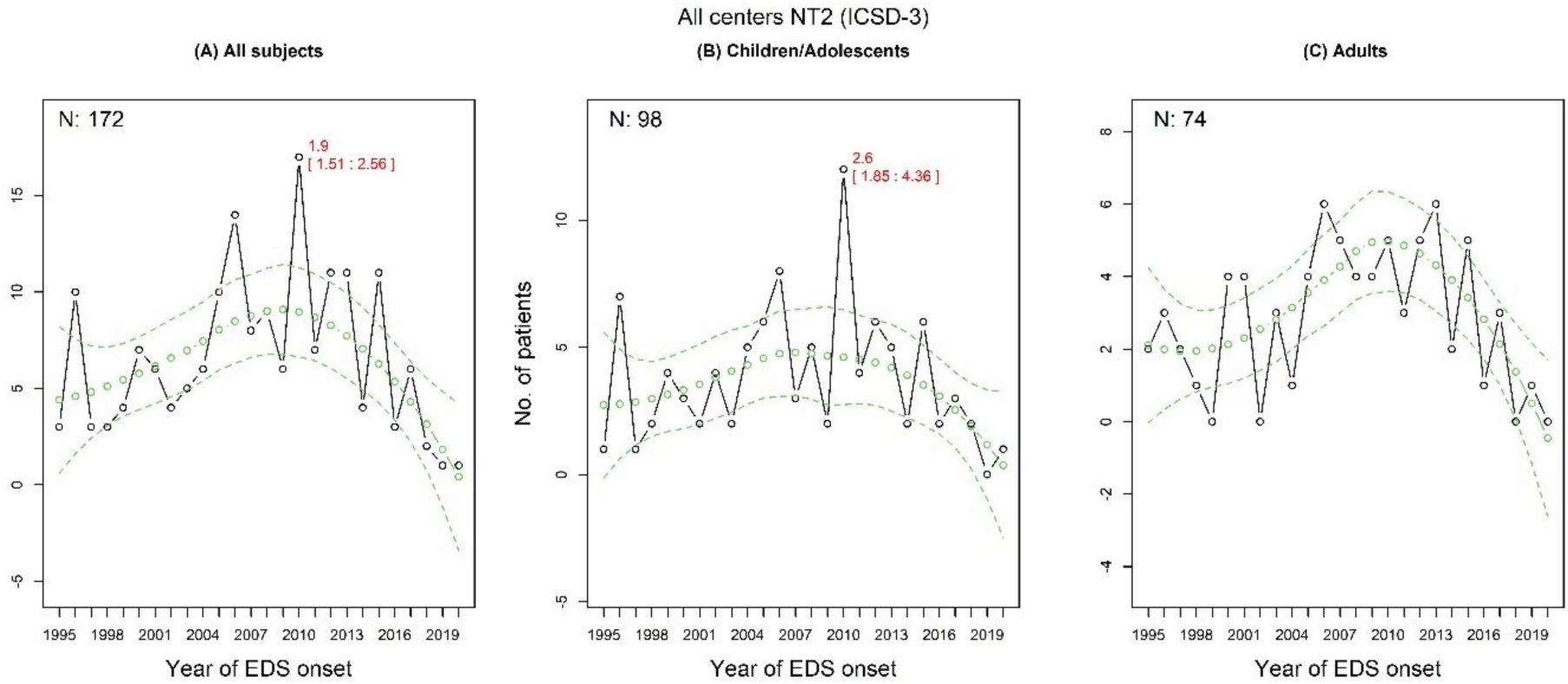
Narcolepsy type 2 incidence rates excluding clinical diagnosis. The changes in narcolepsy type 2 (NT2) in all individuals **(A)**, children **(B)** and adults **(C)**. The predicted incidence rates and their 95% predictive confidence intervals are marked as green circles/lines and the actual values are in black circles. EDS: excessive daytime sleepiness.

**Supplementary figure 14.**
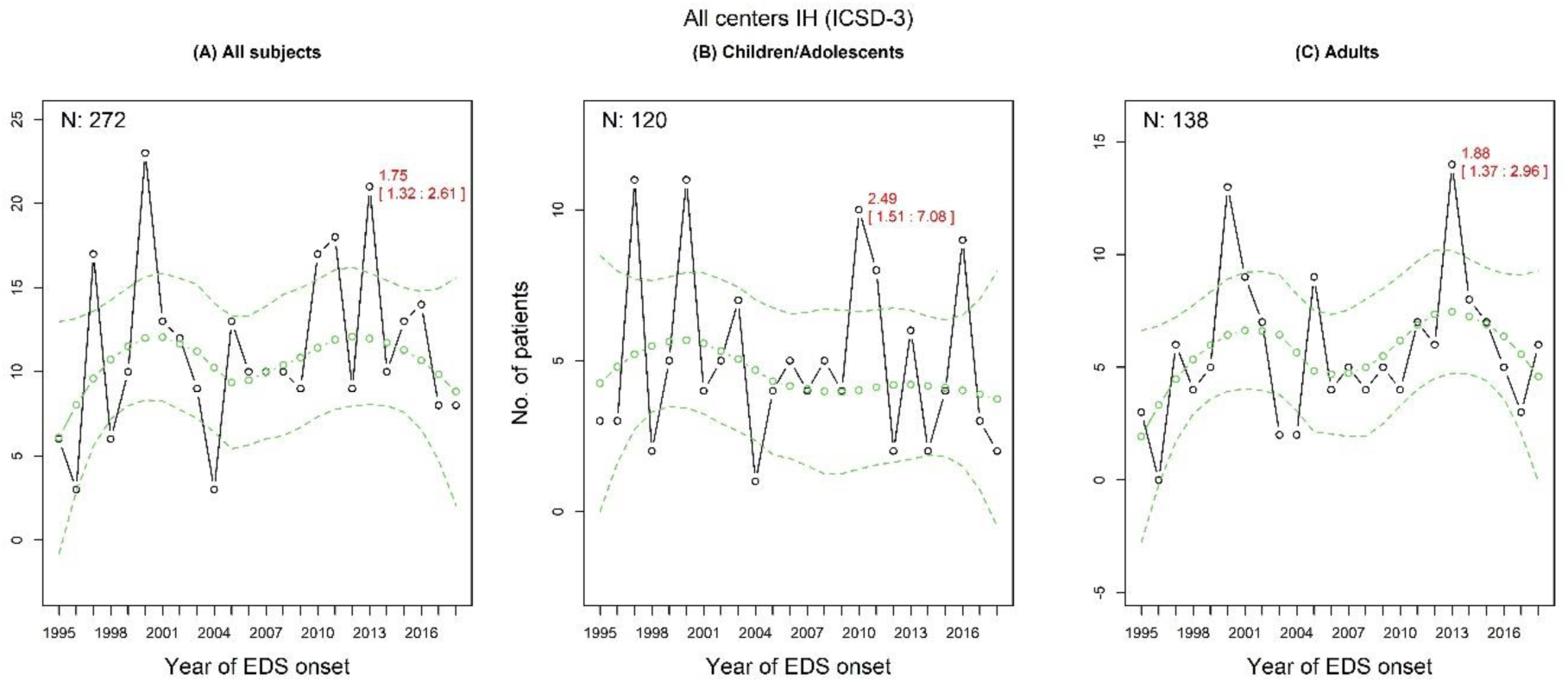
Idiopathic hypersomnia incidence rates excluding clinical diagnosis. The changes in idiopathic hypersomnia (IH) in all individuals **(A)**, children **(B)** and adults **(C)**. The predicted incidence rates and their 95% predictive confidence intervals are marked as green circles/lines and the actual values are in black circles. EDS: excessive daytime sleepiness.

### Narcolepsy type 1 incidence and preceding influenza season severity excluding clinical diagnosis

**Supplementary figure 15.**
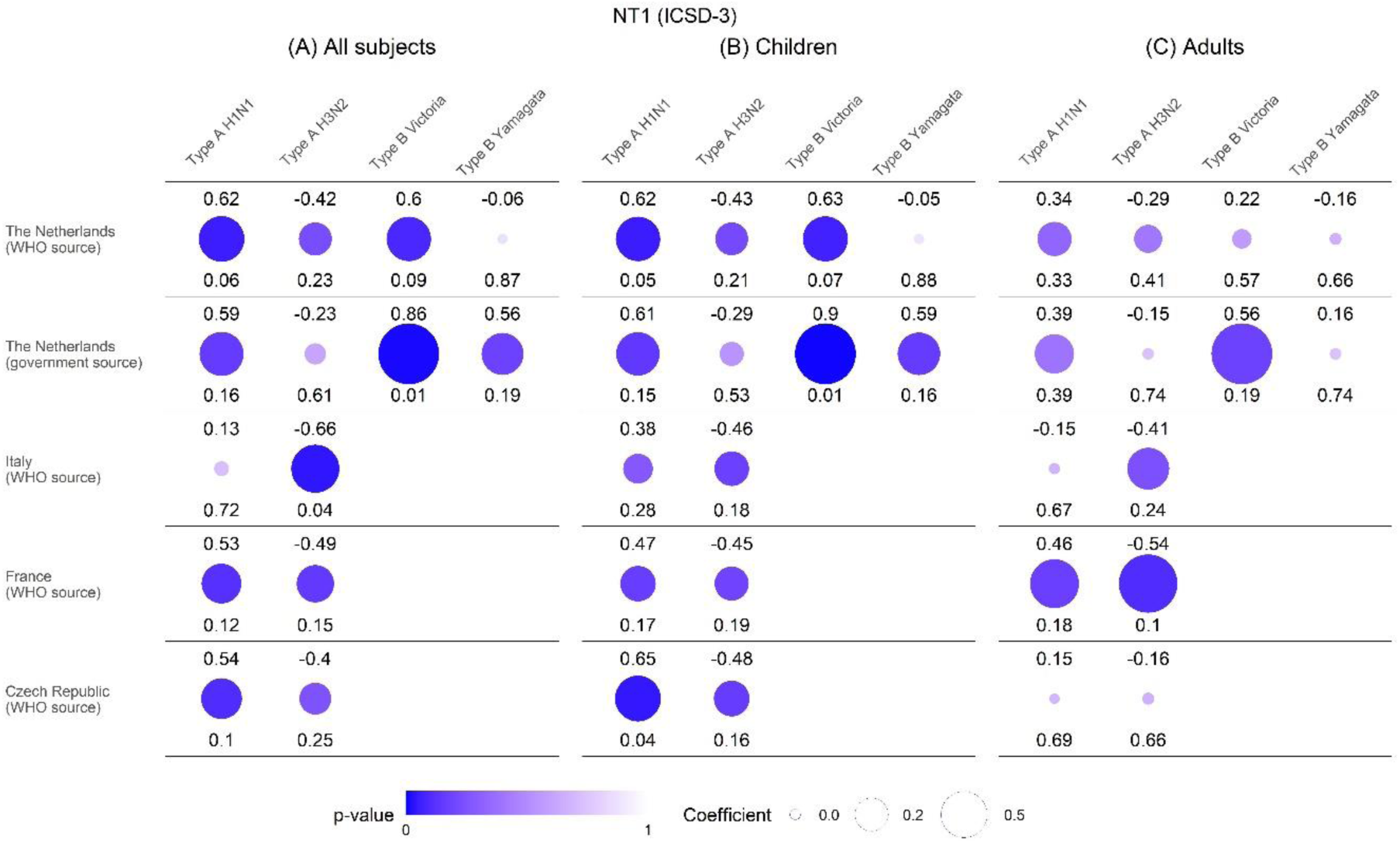
Narcolepsy type 1 and preceding influenza season severity correlations per country excluding clinical diagnosis. The three panels respectively represent the correlations between annual incidence rates of each influenza strain incidence and narcolepsy type 1 (NT1) incidence in the following year for all individuals **(A)**, children **(B)** and adults **(C)**. Top values and circle sizes represent the correlation coefficients. Bottom values and circle colours represent the level of significance of the correlation.

### Sensitivity analysis on narcolepsy type 1 incidence rates and influenza season severity when shifting the influenza season severity one year backward

**Supplementary figure 16.**
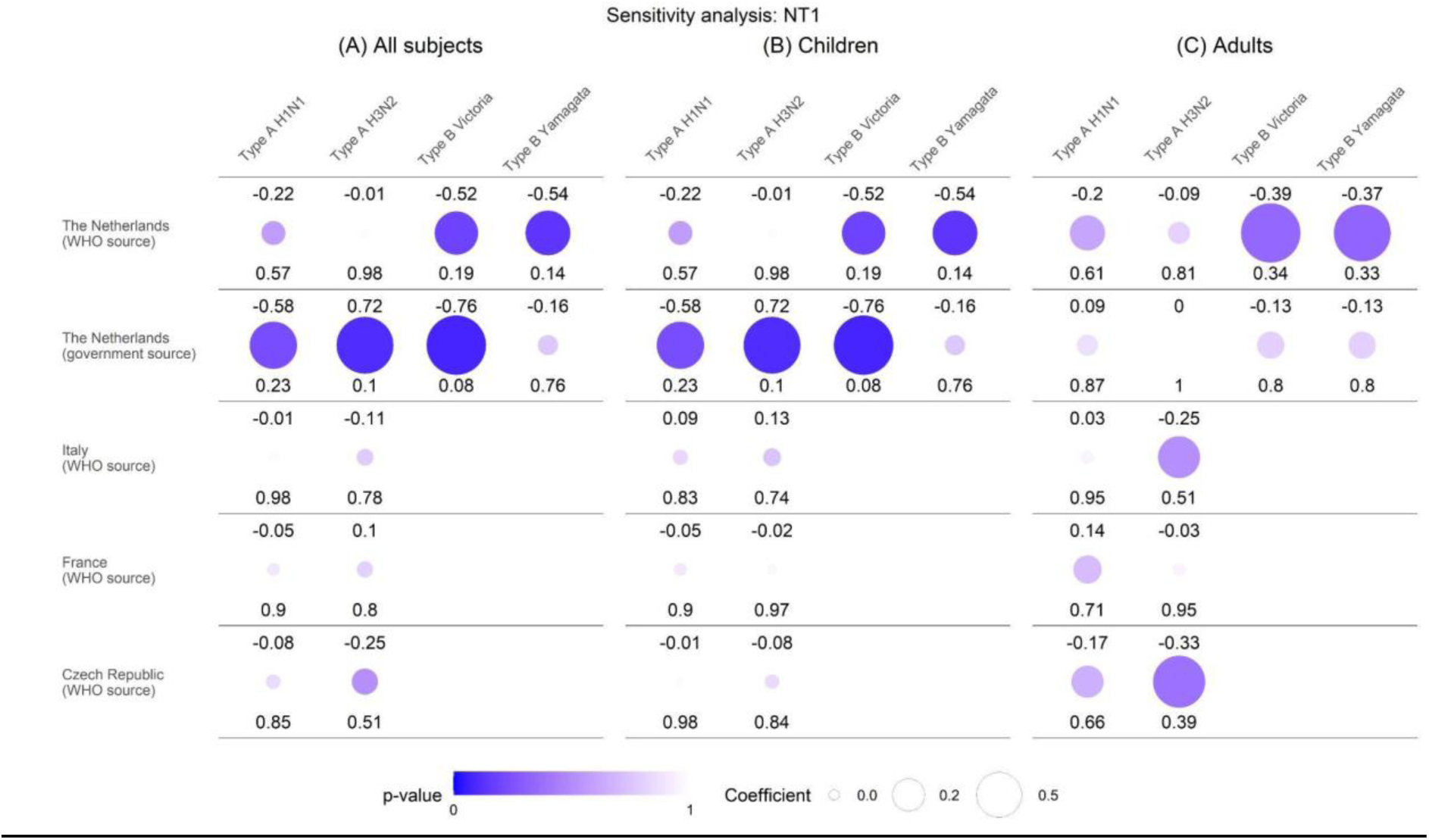
Sensitivity analysis on narcolepsy type 1 and influenza season severity correlations per country when shifting influenza season severity one year backward. The three panels respectively represent the correlations between annual incidence rates of each influenza strain incidence and narcolepsy type 1 (NT1) incidence in the following year for all individuals **(A)**, children **(B)** and adults **(C)**. Top values and circle sizes represent the correlation coefficients. Bottom values and circle colours represent the level of significance of the correlation.

## Notes

### Competing Interest Statement

The authors have declared no competing interest.

### Funding Statement

This study did not receive any funding

